# Mind the gap: characterizing bias due to population mismatch in two-sample Mendelian randomization

**DOI:** 10.1101/2025.07.30.25332465

**Authors:** Jack Li, Jean Morrison

## Abstract

Mendelian randomization (MR) is a statistical method for estimating causal effects using genetic variants as instrumental variables. In two sample MR (2SMR), different study samples are used to estimate genetic associations with the exposure and outcome. For valid inference, these studies must include individuals from the same population. Using studies from different populations may bias the MR estimate due to differences in variant-exposure associations resulting from differences in linkage disequilibrium or genetic effects on the exposure trait. We show that violation of the same-population assumption leads to bias in the causal estimate towards zero on average, and does not increase the rate of false positives when using the most common MR study design. We verify this result in a broad survey of MR estimates, comparing estimates made with matching and mismatching populations across 546 trait pairs measured in 2-7 ancestries. We find that most population-mismatched estimates are attenuated towards zero compared to their corresponding population-matched estimates, and that increasing genetic distance between study populations is associated with greater shrinkage. We observe bias even when mismatched populations have the same continental ancestry. However, we also find that, in some cases, using a larger exposure study with mismatching ancestry can improve power by dramatically increasing precision. These results show that even intra-continental population mismatch can bias MR estimates, but also suggests there is potential to improve the power of MR in understudied populations by properly leveraging larger, mismatching study populations.

## 2 Introduction

Mendelian randomization (MR) is a popular statistical method for estimating the causal effect of an exposure on an outcome using genetic variants as instrumental variables ^1^. MR has become extremely common in recent years due to its potential to estimate causal effects using only summary information for genetic associations, which are readily available through public resources such as the OpenGWAS Project and the EMBL-EBI GWAS catalog ^2;3^. We focus on this summary-level form of MR throughout our analysis. An advantage of summary statistic MR is that it can be conducted using variant-exposure and variant-outcome associations measured in fully separate or partially overlapping samples (two-sample MR; 2SMR). This is the most popular variant of MR because it enables estimation of a wide range of causal effects by not requiring that both exposure and outcome are measured in the same sample. Additionally, using many common estimation methods, 2SMR is preferable to one-sample MR (1SMR) with fully overlapping samples because 2SMR estimates using non-overlapping samples are biased towards the null while 1SMR estimates are biased towards the confounded population association ^4^. Moreover, advanced methods such as GRAPPLE ^5^, CAUSE ^6^, and MR-BEE ^7^ can account for this bias and for partial sample overlap in 2SMR.

Valid MR estimation relies on the assumption that the exposure and outcome GWAS studies are sampled from the same underlying population, known as the same-population assumption. Haycock et al. ^1^ note that even though it may be possible to test the hypothesis of no causal effect when the same-population assumption is violated, the magnitude of the causal estimate may not be accurate. Moreover, Burgess et al. ^8^ note that a variant may be a valid instrument in one population but not the other, and that more generally, an association in one population may not be replicated in another. This observation is strengthened by work finding a genetic correlations less than one for the same trait measured in different populations ^9;10^. Zhao et al. ^11^ show that when there is population mismatch but variant-exposure causal effects are linear and the same across populations (structural invariance), the two-stage least squares estimator is unbiased but inefficient. However, this estimator is biased if either linearity or structural invariance fails. Finally, Woolf et al. ^12^ develop a simple falsification test to validate this same-population assumption by comparing the GWAS effect estimates of variants selected as instruments across populations.

The same population assumption is acknowledged in the Strengthening the Reporting of Observational Studies in Epidemiology using Mendelian Randomization (STROBE-MR) guidelines, which recommend that authors describe the study design and underlying population of each dataset ^13^. Typically, investigators conducting MR studies attempt to fulfill the same-population assumption by selecting GWAS with the same continental ancestry. For example, a multi-ancestry MR study of the effect of the proteome on a common diseases used pQTL data from Europeans from ARIC and INTERVAL and outcome disease GWAS of Europeans from the Global Biobank Meta-Analysis Initiative (GBMI) ^14^. However, GWAS used in MR studies often do not match beyond continental-level ancestry. For example, researchers commonly use summary statistics from meta-analyses in MR studies due to their large sample sizes ^15;16;17;18;19^. This practice guarantees some amount of population mismatch between exposure and outcome samples.

While the theoretical basis of the same population assumption is well understood, the magnitude of bias due to violations in common real-world MR designs is unknown. Although matching study populations on continental ancestry is common, it is unclear how effective this practice is at mitigating bias due to violation of the same-population assumption. Additionally, it is possible that this practice is too restrictive and inhibits performing MR studies of effects in populations for which there are few or under-powered population-matched exposure GWAS. It may also discourage researchers from making full use of large multi-ancestry meta analyses, potentially reducing power of the resulting analyses.

Studies by Wang et al. ^10^ and Ding et al. ^20^ have examined the portability of polygenic risk scores between different ancestries, finding that attempting to use mismatching populations leads to reduced prediction accuracy due to differences in LD patterns and MAF ^10^, with reduced accuracy even between individuals in the same genetic ancestry clusters ^20^. However, analogous literature quantifying the impact of population mismatch in MR across a broad range of possible traits, ancestries and causal effects is sparse. In this paper, we characterize how violations of the same-population assumption affect MR estimation. First, we show theoretically that violations of the same-population assumption are expected to bias the MR causal effect estimate towards zero using the most common instrument selection strategy. Second, we compare population-matched and population-mismatched MR estimates across a wide range of traits and ancestries to systematically estimate bias due to population differences. In agreement with our theoretical expectations, we find that population mismatch nearly always attenuates signals of causal effects toward zero. Finally, we evaluate the trade-off between bias and power when choosing between a smaller population-matched exposure study and a large population-mismatched exposure study. Our results suggest that, in some cases, substantial power gain can be realized by violating the same-population assumption, at the cost of null-biased effect estimates.

## 3 Methods

### 3.1 Effect of Population Mismatch on MR Estimates

#### 3.1.1 Statistical Setting and Assumptions

We first assume that exposure *X* and outcome *Y* are related by the linear structural equation *Y*_*i,t*_ = *γ*_*t*_*X*_*i,t*_+*v*_*i,p*_, for individual *i* in the target population, *t*. In our setting, the causal effect of *X* on *Y* in population *t, γ*_*t*_, is the target of inference. We assume that GWAS estimates of the marginal linear associations of variants *G*_1_, …, *G*_*J*_ with *Y* are measured in population *t* and estimates of the marginal linear associations of the same set of variants with *X* are measured in population *e*, the “exposure population”. Populations *e* and *t* may or may not differ.

If populations *e* and *t* are the same, consistent estimation of *γ*_*t*_ using inverse-variance weighted (IVW) regression and similar summary statistic MR methods requires four assumptions. The first three are commonly designated as “valid IV assumptions” and are (i) *G*_*j*_ is associated with *X*, (ii) there are no confounders of *G*_*j*_ and the outcome, and (iii) *G*_*j*_ is only associated with *Y* through its association with *X* ^21^. Assumption (i) requires that *G*_*j*_ and *X* are not independent. However, in the case of summary statistic MR, we require a more stringent version of this assumption, that the expectation of the regression coefficient regressing *G*_*j*_ on *X* is not zero. Assumptions (ii) and (iii) are satisfied by the condition that *G*_*j*_ is independent of *Y* given *X*. Finally, a fourth assumption is required for identifiability of the causal effect of *X* on *Y*. One common version of this assumption is (iv) homogeneity of the *G* − *X* association, meaning that the linear association of *G*_*j*_ with *X* does not vary across individuals. Weaker variations of the fourth assumption, including no simultaneous heterogeneity (NOSH) ^22^ and monotonicity ^23^ are discussed in Supplemental Note Section 1.1. If assumptions (i)-(iii) and at least one variation of assumption (iv) hold in population *t*, then

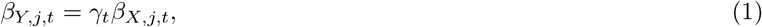

where *β*_*Y,j,t*_ is the average marginal association of *G*_*j*_ with *Y* and *β*_*X,j,t*_ is the average marginal association of *G*_*j*_ with *X*, both in population *t*. All summary statistic MR methods rely either on (1) or an extension of (1) to *β*_*Y,j,t*_ = *γ*_*t*_*β*_*X,j,t*_ +*α*_*j*_. The term *α*_*j*_ captures associations between *G*_*j*_ and *Y* not mediated by *X* (violations of assumption (iii)), and is usually assumed to follow a known distributional form. IVW regression ^24^ as well as estimators such as the weighted median ^25^ estimate *γ*_*t*_ as a weighted average of the ratios 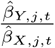. Other methods such as GRAPPLE ^5^ or CAUSE ^6^ are likelihood-based, but still rely on equation (1) or its extension including *α*_*j*_.

When populations *e* and *t* differ, a fifth assumption, (v) that *β*_*X,j,e*_ = *β*_*X,j,t*_ for all *j*, is additionally required. This assumption ensures that *β*_*Y,j,t*_ = *γ*_*t*_*β*_*X,j,e*_. If this assumption holds, summary statistic MR estimators consistently estimate *γ*_*t*_, the causal effect in the population in which variant-outcome associations are measured. We note that we have not made any assumptions about the causal effect of *X* on *Y* in the exposure population. In our setting, no information about *Y* in population *e* is available, so the relationship between *X* and *Y* in population *e* does not impact the MR estimate. Therefore, we always interpret the MR estimate as the causal effect of *X* on *Y*, specific to the target (outcome) population ^11^. Further discussion of interpretation of the MR estimate when the *X* − *Y* relationship is nonlinear or heterogeneous is given in Supplemental Note Section 1.1.

#### 3.1.2 Typical IV selection practices induce shrinkage in MR with mismatching populations

Figure 1 illustrates the design of a summary-statistic MR study, as well as the components of standard summary-level MR estimates. Under an additive genetic model, the true association of *G*_*j*_ with *X* can be expressed as a weighted combination of the causal effects of variants in linkage disequilibrium (LD) with *G*_*j*_ on *X*,

**Figure 1.**
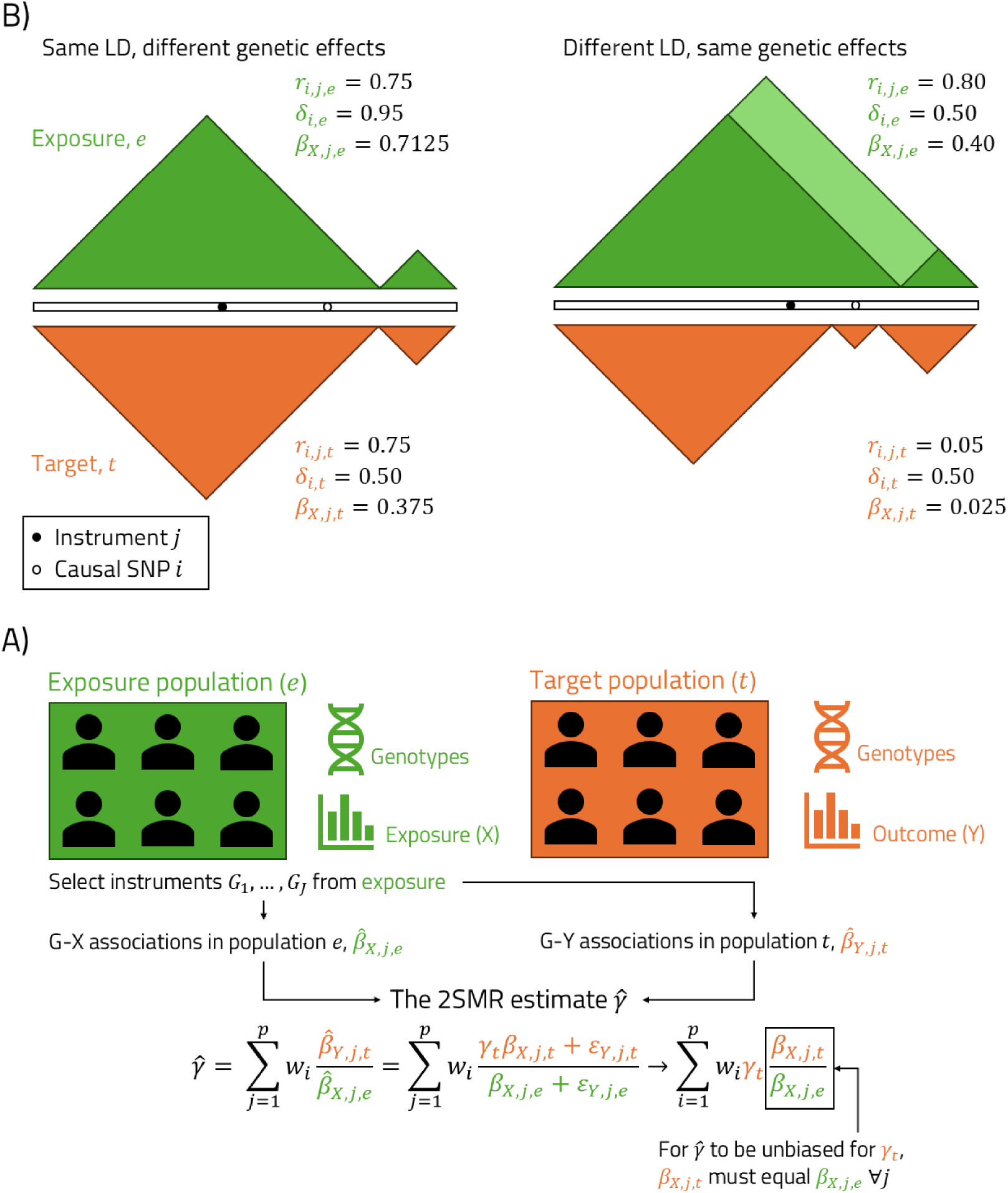
(A) Diagram depicting a 2SMR study and the calculation of the causal estimate as a weighted average of ratio estimates. (B) Two possible sources of differences between *β*_*X,j,t*_ and *β*_*X,j,e*_. Left, LD patterns are the same in the two populations but the causal variant has a different effect on the exposure. Right, the causal variant has the same effect in both populations, but LD structure differs. In both cases, there is a strong exposure-instrument association in population *e*, but a weaker association in population *t*.

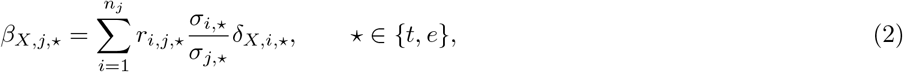

where *δ*_*X,i*,⋆_ is the average per-allele effect of variant *i* on *X* in population ⋆, *r*_*i,j*,⋆_ is the correlation between variant *i* and variant *j* in population ⋆, and *σ*_*i*,⋆_ and *σ*_*j*,⋆_ are genotype standard deviations calculated by 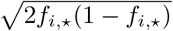 and 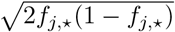, where *f*_*i*,⋆_, *f*_*j*,⋆_ are the allele frequencies of variants *i* and *j* in population ⋆. This sum is over all variants in LD with variant *j* ^26^. Thus, if any of LD, allele frequencies, or causal genetic effects differ across populations, then *β*_*X,j,e*_ is not equal to *β*_*X,j,t*_ and assumption (v) fails.

Population differences in *r*_*i,j*,⋆_, *f*_*i*,⋆_ and *f*_*j*,⋆_ can occur due to differences underlying recombination rates, selection, mutation rates, or by chance. In general, we expect these differences to be larger between pairs of populations with larger fixation index (*F*_*st*_). Differences between *δ*_*X,i,e*_ and *δ*_*X,i,t*_ can occur due to gene-environment or gene-gene interactions, or due to nonlinear casual genetic effects in combination with differences in allele frequency ^11^. Differences in these three factors are also responsible for the imperfect transferability of polygenic scores between populations ^10;20^.

When assumption (v) fails, *β*_*X,j,e*_ could differ from *β*_*X,j,t*_ in any direction. However, in most MR studies, variants are selected as instruments based on their strength of association with *X* in the exposure population (Figure 1). This process induces survivorship bias. A variant that is strongly associated with *X* in the target population but weakly associated in the exposure population is much less likely to be selected than a variant that is strongly associated with *X* in the exposure population and weakly associated in the target population. This instrument selection practice means that, on average, |*β*_*X,j,e*_| will be larger than |*β*_*X,j,t*_| for selected variants, under reasonable conditions on the distribution of causal effects (see Supplemental Note Section 1.2). This survivorship bias is similar to the well-documented phenomenon of winner’s curse in GWAS. Xiao and Boehnke ^27^ observe that GWAS associations are typically smaller in magnitude in a replication cohort than a discovery cohort, and Palmer and Pe’er ^28^ note that this effect is stronger when discovery and replication cohorts have different ancestries. As a result, 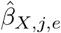 is, on average, an overestimate of *β*_*X,j,t*_, meaning that the summary statistic MR estimate made with mismatching populations will have an expectation that is closer to zero than the estimate made with matching populations. In the absence of a true causal effect in the target population, *β*_*Y,j,t*_ is equal to zero as long as the selected variant does not exhibit horizontal pleiotropy in the target population. This means that summary statistic MR estimators are unbiased under the null hypothesis if the first four MR assumptions are met, even if assumption (v) fails.

Horizontal pleiotropy, or failure of assumption (iii), is common in MR studies and results in bias when using methods that do not appropriately model failure of this assumption. In particular, this bias can result in an increased risk for false positives. However, the presence or magnitude of horizontal pleiotropy is independent of the probability that a variant is selected as an instrument, regardless of which population is used for selection. This is because variants are selected based only on their association with *X*, and not with *Y*. Therefore, we do not expect violation of the same-population assumption to increase the risk of false positives due to horizontal pleiotropy.

### 3.2 Empirical Survey of the Impact of Population Mismatch on MR Estimates

#### 3.2.1 Study Design

To empirically assess the effect of population mismatch on MR estimates, we compared MR estimates made in matching populations to those made in mismatching populations across a large number of traits and populations, as illustrated in Figure 2. We selected 26 exposure traits including anthropometric, clinical, and molecular measurements. For each exposure trait, we identified one outcome trait with prior evidence of a causal link, resulting in a total of 21 outcome traits, as some exposures were linked to the same outcome. These include 20 binary diseases and one continuous trait, total billirubin. This strategy ensures that the set of all pairwise combinations of exposure and outcome traits includes some expected non-null effects as well as some likely null effects. Exposure and outcome traits are listed in Supplemental Note Tables S1 and S2.

**Figure 2.**
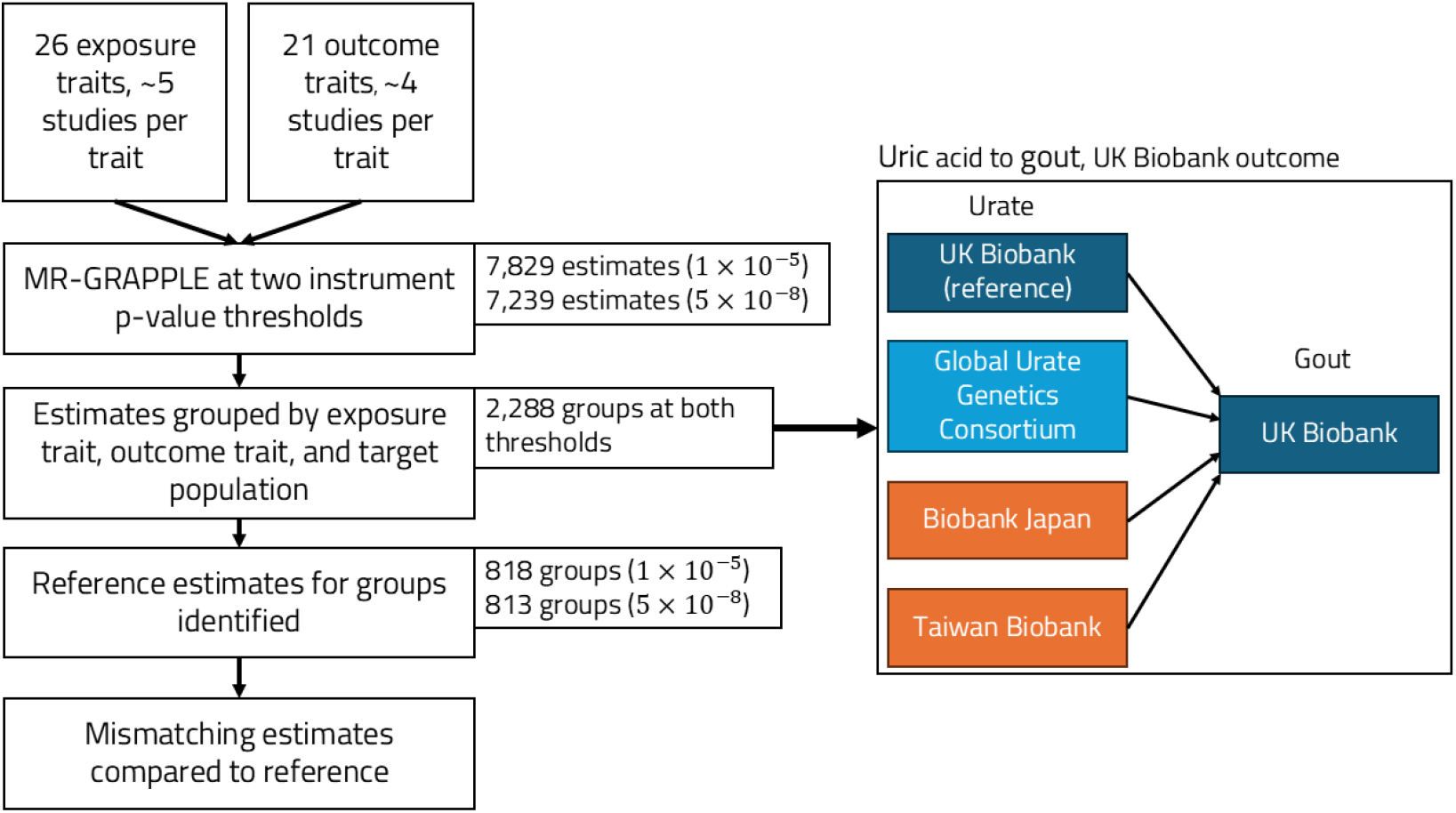
Study design of cross-population survey of MR estimates. Right, example of an exposure trait-outcome study group for the effect of urate on gout with UK Biobank defining the target population. Exposure studies are color-coded based on how closely study populations match the target population: exact match (dark blue), continental match but a different subpopulation (light blue), or a continental mismatch (orange).

We identified publicly available GWAS summary statistics for all exposure and outcome studies from as many populations as possible using the OpenGWAS database, the EMBL-EBI GWAS catalog, and FinnGen Release 10^2;3;29^. We identified an average of 5 studies per exposure trait (128 total exposure studies) and 4 studies per outcome trait (88 total outcome studies). In total, there are 546 exposure-outcome trait pairs, and 11,392 exposure-outcome study pairs (approximately 21 study pairs per trait pair). The study source for each trait is given in Supplemental Data 1.

MR estimates the causal effect in the target population, which is the population in which the outcome GWAS is performed. Therefore, we compared MR estimates within 88 *×* 26 = 2, 288 groups defined by an exposure-outcome pair and a target population (Figure 2). This ensures that any differences observed in MR estimates are due to differences in the exposure population and not population differences in the causal effect.

Different studies of the same exposure may report effects in different units. In order to obtain comparable MR effect size estimates from different exposure studies, we converted all exposure studies for the same trait to a standard unit, such as mmol/L for LDL cholesterol. In the case of studies that were inverse-normalized or z-score transformed, we scaled variant effect estimates by the standard deviation of the exposure trait if it was listed, or used an external estimate when feasible. Studies in which the exposure trait was nonlinearly transformed (e.g., log or square root) and meta-analyses that combine estimates on different scales could not be converted to a standard unit but were retained for comparisons of sign and significance of causal estimates. In total, we were able to convert 89 of the 128 exposure studies to a standard unit (Supplemental Data 2). Estimates for binary outcome were all either reported on the log odds scale or converted from the standardized scale to the log odds scale by multiplying by *µ*(1 − *µ*), where *µ* is the proportion of cases in the study cohort (Supplemental Data 3).

#### 3.2.2 Mendelian Randomization

We computed MR estimates for all exposure-outcome trait and study pairs using IVW regression, GRAPPLE ^5^, and MRBEE ^7^. For all three estimators, instruments were selected from the exposure GWAS after LD clumping, prioritizing variants with the strongest exposure association with threshold *r*^2^ *<* 0.01. This practice can result in a small amount of bias due to winner’s curse ^21^ but is the most common approach used in 2SMR studies. For IVW regression, instruments were selected using a p-value threshold of 5 *×* 10^−8^. For GRAPPLE and MRBEE, we used two thresholds, 5 *×* 10^−8^ and 1 *×* 10^−5^ because these methods are more robust to weak instrument bias. We additionally performed Steiger filtering to remove variants more strongly associated with the outcome than the exposure.

We computed each estimator for all exposure-outcome study pairs with at least two instruments. Using 5 *×* 10^−8^ as the instrument selection p-value threshold, we could analyze 9, 828 pairs, of which 7,239 are on a standard scale. Using the 1 *×* 10^−5^ threshold, we could analyze 11, 248 pairs, of which 7,829 are on a standard scale. Complete details of our implementation of Steiger filtering and both MR estimation methods are given in Supplemental Note Section 2.

We compared MR estimates made in matching and mismatching populations, under the assumption that differences in these estimates are due to violation of the same-population assumption. However, this assumption is not always justifiable using the IVW regression method, due to sample overlap in the population-matched analysis. When MR is performed in overlapping samples, weak-instrument bias in IVW regression is towards the observational association, rather than towards zero ^4^. GRAPPLE and MRBEE estimates do not suffer from this problem because sample overlap is accounted for using a pre-computed estimate of the residual correlation between variant-exposure and variant-outcome association estimates ^5^. Differences in IVW regression estimates could also occur due to differences in instrument strength between two different exposure studies resulting in different magnitudes of weak instrument bias. By using an unbiased estimating function, MRBEE does not suffer from weak instrument bias at all ^7^, while instrument strength has only a small effect on GRAPPLE estimates ^5^. However, GRAPPLE uses a more flexible model for horizontal pleiotropy than MRBEE^5^. Therefore, we present results from GRAPPLE as our primary analysis and include results using MRBEE and IVW regression in the Supplemental Note.

#### 3.2.3 Quantifying the effect of population mismatch on MR Estimates

We define an estimate as population-matched if the exposure and outcome study are sampled from the same study cohort. Of the 2,288 estimate groups defined by an exposure-outcome trait pair and a target population (see Section 3.2.1), 818 (1 *×* 10^−5^ selection threshold) and 813 (5 *×* 10^−8^ threshold) contain a population-matched estimate that can be converted to a standardized scale. For each of these groups, we designate the MR estimate using the matching exposure population as the reference estimate, 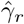.

We used two strategies to quantify bias due to violation of the same-population assumption. First, for each population-mismatched exposure study, we computed a z-score to test the hypothesis that the population-mismatched and reference estimate have the same expectation,

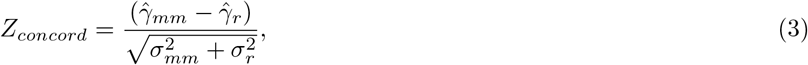

where 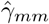 is the population-mismatched estimate, and 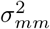 and 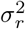 are estimated standard errors of 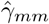and 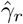. If 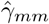 and 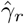 have the same expectation, then *Z*_*concord*_ follows a standard normal distribution. For each z-score we computed a p-value testing the hypothesis that the z-score was drawn from the null distribution. We used the Benjamini-Hochberg procedure to compute estimates of the false discovery rate (FDR) within estimate groups for each instrument selection threshold, using an FDR of 5%. We classified mismatching estimates that were significantly different from their reference estimate at FDR *<* 0.05 into three categories: estimates with the same sign as the reference estimate and closer to zero, estimates with the same sign as the reference estimate and farther from zero, and estimates with the opposite sign as the reference estimate.

Second, to quantify the average degree of shrinkage or inflation resulting from population mismatch, we regressed mis-matching estimates on corresponding reference estimates with no intercept, weighted by the inverse-variance of the mismatching estimates. We used simulation extrapolation (SIMEX) to account for uncertainty in the MR estimates (Supplemental Note Section 3) ^30^. The resulting estimated slope provides an estimate of the expected shrinkage or inflation due to violation of the same-population assumption. An estimate less than 1 indicates that estimates obtained using mismatching populations tend to be closer to zero than estimates obtained using matching populations. We used bootstrapping with 1,000 replicates to estimate the standard error of these SIMEX-estimated shrinkage coefficients.

We computed the SIMEX regression slope across all analyses with available reference estimates and within groups defined by exposure-outcome population pairs. At an instrument selection p-value threshold of 1 *×* 10^−5^, we identified 26 population pairs with at least eight estimates with an available reference estimate. We selected this threshold to ensure that we could calculate SIMEX regression slopes for at least some population pairs using African populations as a target, while maximizing the precision of the estimates we calculate. At the stricter selection p-value threshold of 5 *×* 10^−8^, 23 population pairs have at least eight estimates with an available reference estimate. However, none of these pairs include African populations as a target population. We expect greater shrinkage in population pairs with larger differences in LD. To assess this prediction, we regressed the SIMEX estimated slope against the approximate *F*_*st*_ distance between the populations, obtained from Scott et al. ^31^ (Supplemental Note Table S3), weighted by the inverse of the variance of each estimate. The SIMEX regression procedure does not account for correlation between estimates that share the same exposure or outcome studies. This correlation may lead to under-estimation of standard errors, motivating our use of the bootstrap to estimate standard errors of shrinkage coefficients.

### 3.3 Trade-off between bias and power

To assess the trade-off between bias and power when using a larger mismatching exposure study, we examined 195 causal estimates made using outcomes measured in Biobank Japan. We compared population-matched estimates with estimates obtained using exposures from a meta analysis of UK Biobank and Biobank Japan ^32^, which is nearly three times as large as Biobank Japan alone. As the meta-analysis study combined effects of two populations measured on different scales, the population-mismatched MR estimates computed using the meta-analysis as exposure could not be converted to the standard scale. Therefore, we focus primarily on the sign and significance of these effect estimates.

We identified trait pairs that were significant in the population-mismatched analysis but non-significant in the population-matched analysis and vice-versa using a Bonferroni corrected signficance threshold of *p <* 0.05*/*195. We assessed the replicability of effects found in only one of the two analyses by comparing results with estimates from population-matched analyses using samples that do not overlap with either UK Biobank or Biobank Japan. The significance threshold for replication was set at the Bonferroni adjusted 0.05 threshold accounting for the total number of effects we attempted to replicate.

## 4 Results

### 4.1 Using a mismatching exposure biases MR estimates towards zero

Figure 3 shows GRAPPLE MR estimates for the effect of LDL cholesterol on coronary heart disease in four target populations. Reference estimates are available for Biobank Japan and UK Biobank target populations. There are no reference estimates when using FinnGen or the CARDioGRAMplusC4D studies to measure the outcome, but there are estimates made using exposure studies with majority matching continental ancestry. MR estimates made using mismatching populations are all attenuated towards zero compared to the reference or continentally matched estimates. Using the instrument selection p-value threshold of 1 *×* 10^−5^, all population-mismatched estimates are significantly closer to zero than the reference estimates at an FDR threshold of 5% based on the concordance z-score. We observe the same pattern at the stricter selection threshold, though differences are less significant due to higher variance in the causal effect estimates.

**Figure 3.**
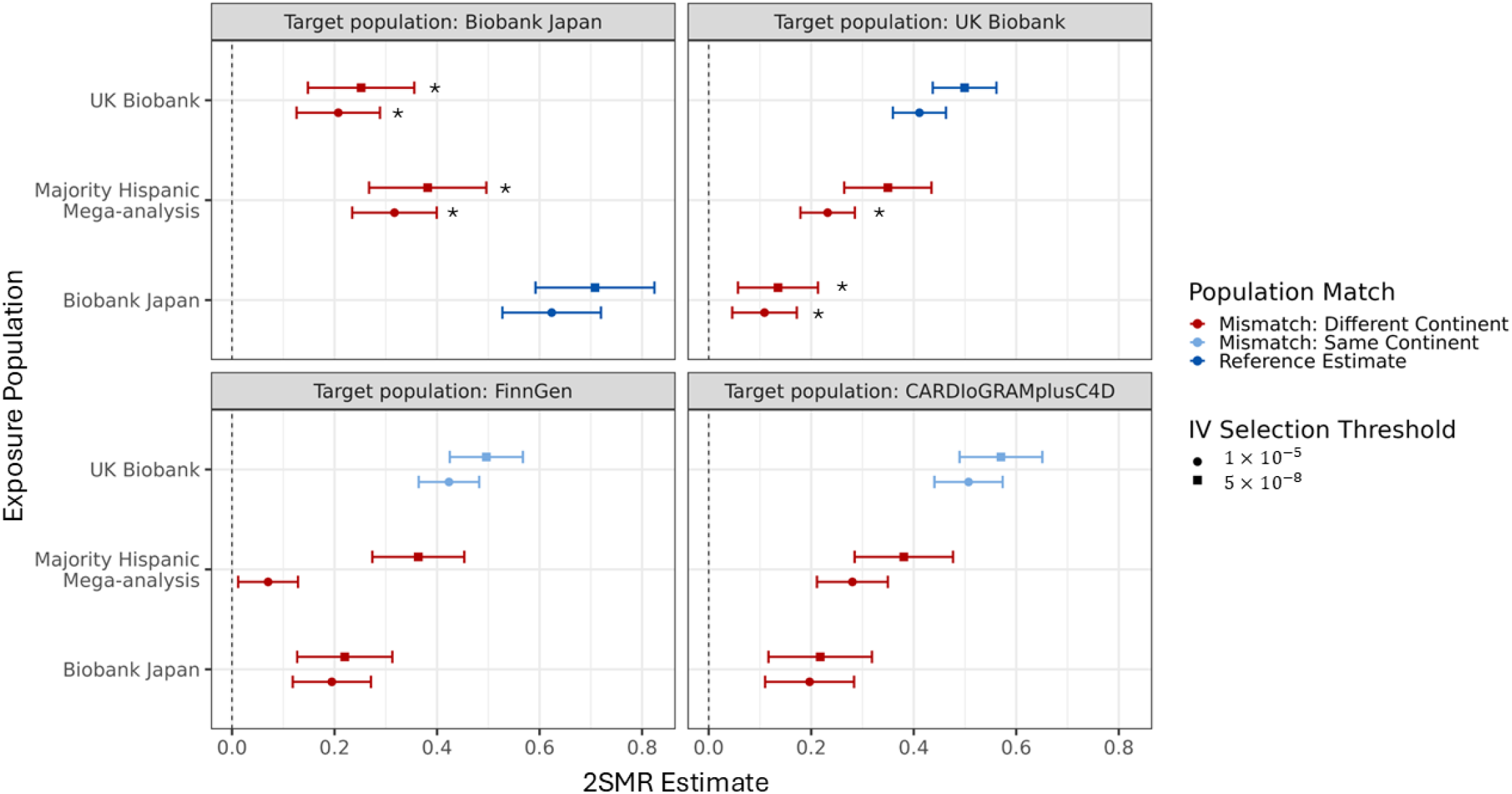
Forest plots of MR estimates of the effect of LDL cholesterol on coronary heart disease from GRAPPLE at two instrument selection p-value thresholds. Points indicate effect estimates and intervals indicate 95% confidence intervals. Color indicates if an estimate is made with exact matching (dark blue), continentally matching (light blue), or continentally mismatching (red) exposure studies. A star indicates that an estimate is significantly different than the reference estimate (FDR *<* 0.05), when a reference estimate is available.

The shrinkage shown in Figure 3 is exemplary of a general pattern across all trait pairs. The SIMEX-estimated overall shrinkage coefficient is significantly less than 1 at both instrument selection thresholds (0.41 (SE 0.015) for 1 *×* 10^−5^ threshold and 0.46 (SE 0.019) for 5 *×* 10^−8^ threshold). At both instrument selection thresholds, the majority of population-mismatched estimates with significant concordance z-scores were of the same sign and closer to zero than their corresponding reference estimates (285/404 (71%) for 10^−5^ threshold, 105/160 (66%) for 5 × 10^−8^ threshold). In the majority of the remaining cases, the population-mismatched estimate has opposite sign from the reference estimate. However, in most of these cases (113/118 (96%) for 10^−5^ threshold, 43/50 (86%) for 5 *×* 10^−8^ threshold) either the population-mismatched or reference estimate is not significantly different from zero at a nominal 0.05 significance threshold. We found only one (10^−5^ threshold) and five (5 *×* 10^−8^ threshold) instances of a significant concordance z-score and a population-mismatched estimate that is of the same size and larger in magnitude than the corresponding reference estimate (Supplemental Note Table S4). We found similar results using both IVW regression and MRBEE (Supplemental Note Table S5). Recapitulation of results using MRBEE confirms that the observed attenuation of effect sizes is not an artifact of differences in instrument strength. IVW regression reference estimates may be biased due to sample overlap and weak-instrument bias. However, these are provided because IVW regression is one of the most commonly used MR methods.

Full details of all 7,829 and 7,239 standardized MR estimates with and without Steiger filtering at both instrument selection thresholds using GRAPPLE and at the stringent selection threshold using IVW are provided in Supplemental Data 5, 6, and 7. The use of Steiger filtering did not substantially impact most MR estimates (Supplemental Note Figure S1) or the main findings of this section (Supplemental Table S6).

### 4.2 Average shrinkage due to population-mismatch is correlated with *Fst*

Figure 4 shows population pair-specific SIMEX regression estimates (shrinkage coefficients) vs approximate *F*_*st*_ distance between populations using the selection threshold 1*×*10^−5^ results. We observe a clear negative relationship between regression estimates and *F*_*st*_, supported by inverse-variance weighted regression of shrinkage coefficients against *F*_*st*_, which gives a regression coefficient of −2.40 (SE 0.56, p-value 0.0004). However, we also note that *F*_*st*_ does not completely determine the amount of shrinkage for all population pairs and there is a lack of symmetry in shrinkage coefficients between population pairs. For example, the shrinkage coefficient for UK Biobank used as exposure and FinnGen defining the target population is 0.67 (SE 0.03), while measuring the exposure in FinnGen and outcome in UK Biobank results in a shrinkage coefficient of 0.52 (SE 0.02). Notably, the shrinkage coefficient is significantly less than one for all population pairs, even those with matching continental ancestry (Supplemental Note Figure S2). Results are similar using the 5 *×* 10^−8^ variant selection threshold, though these do not include pairs involving African populations (Supplemental Note Figure S2 and Supplemental Note Section 4).

**Figure 4.**
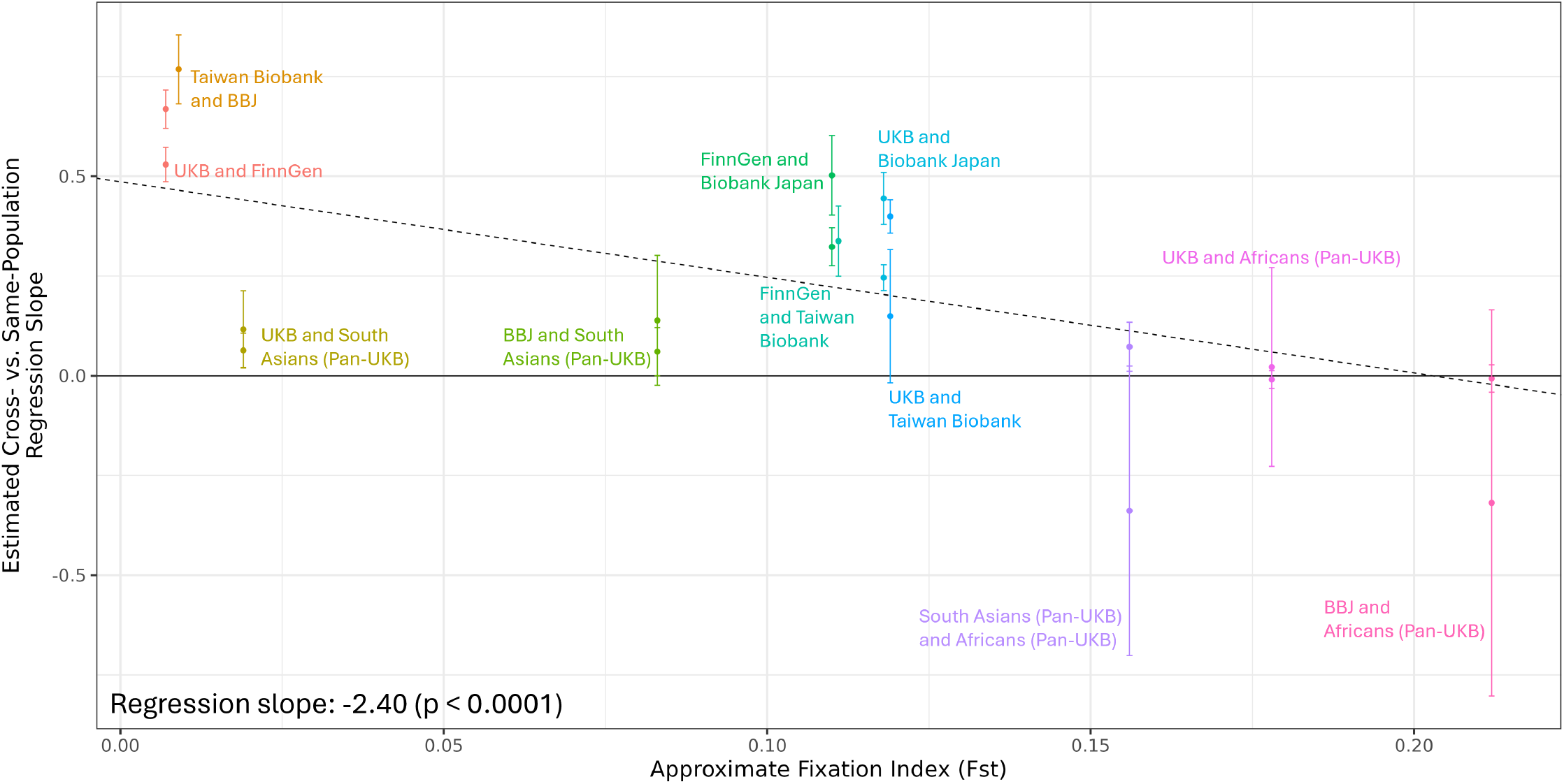
Population pair specific shrinkage coefficients vs approximate *Fst* distance between populations. Population pair specific shrinkage coefficients were estimated using SIMEX. The dashed line represents the univariable regression line from inverse-variance weighted regression of *F*_*st*_ on estimated shrinkage coefficient. Vertical lines represent 95% confidence intervals for each shrinkage coefficient. Colors denote population pair labeled on the plot.

### 4.3 Tradeoff between bias and power

Of the 195 causal estimates computed using Biobank Japan as both exposure and outcome population, 13 are significantly different from zero at *p <* 0.05*/*195 using the more stringent instrument selection threshold, and 17 are significantly different from zero using the more lenient threshold. By contrast, when using a meta analysis of UK Biobank and Biobank Japan as the exposure study, 25 and 26 estimates are significantly different from zero at the more and less stringent instrument selection thresholds, respectively. Using the larger exposure study results in 14 significant causal effects that are not discovered using the population-matched exposure study at both instrument selection thresholds, while 2 and 5 causal estimates are significant in the population-matched analysis only at the more and less stringent instrument selection thresholds, respectively (Supplemental Note Tables S7 and S8 and Supplemental Data 8).

We checked if each causal estimate that differed in significance in the population-matched and population-mismatched analyses replicated in population-matched estimates for non-UK and non-Japanese populations. We were able to perform replication analysis on 13 of the 19 causal effects that differed in significance at the 1 *×* 10^−5^ threshold, and 12 of the 16 trait pairs that differed in significance at the 5 *×* 10^−8^ threshold. At both instrument selection thresholds, all estimates that were significant only using BBJ alone as the exposure study replicated in at least one other population. At the more lenient instrument selection threshold, 8 out of 10 trait pairs identified as significant only using the meta-analysis as the exposure study exposure showed evidence of replication (Supplemental Note Table S7). At the more stringent selection threshold, only 5 out of the 11 trait pairs identified as significant only using the meta analysis showed evidence of replication. However, five out of six of these non-replicating causal effects at the more stringent threshold only had estimates from studies with fewer than 50,000 participants available for replication analysis (Supplemental Note Table S8 and Supplemental Data 9).

## 5 Discussion

We demonstrate, both theoretically and through a comprehensive empirical survey, that bias in MR estimates due to population mismatch usually results in a reduction in magnitude of the estimated effect. This bias occurs even when using continentally matching populations, a common practice in existing MR literature. The mechanism for this bias is non-deterministic, and bias away from zero may occasionally occur. However, over approximately 2,000 comparisons, we found fewer than a dozen instances where population-mismatched estimates were significantly different from their reference estimate and either larger in magnitude or of opposite sign with both estimates nominally significantly different from zero. This suggests that while possible, anti-conservative bias due to population is rare. The observed shrinkage towards zero in population-mismatched estimates is a result of selecting variants as instruments based on strong associations with the exposure in the exposure population. This is the most common procedure used to select instruments in MR. Alternative selection procedures may not necessarily produce nullward bias.

Additionally, we demonstrate a bias-variance tradeoff between population mismatch and sample size using the example of outcomes measured in Biobank Japan and exposure studies sourced either from Biobank Japan or from a meta analysis of UK Biobank and Biobank Japan. We show that using a larger but population-mismatched exposure study may improve power compared to a smaller, population-matched study, at the expense of some bias towards zero in causal estimates. We believe this increase in power is due to the increased precision of estimation due to a large sample size. Larger sample size results in more variants with significant associations with the exposure and more precise estimates of variant-exposure associations. By leveraging LD and allele frequency structure from many different populations, multi-ancestry meta-analyses can increase the resolution of fine-mapping and help identify ancestry-specific causal variants ^33^. Thus, using meta-analyses as exposures in MR may help identify instruments that are closer in LD to the causal variant across all populations or are themselves causal variants, improving their transferability and reducing the bias in MR induced by population mismatch. Moreover, we argue theoretically that estimates made with population-mismatched studies are not more likely to be false positives than those made using matching studies, though both may be subject to bias due to horizontal pleiotropy if this is not appropriately accounted for. This suggests that there may be utility in violating the same-population assumption in MR analysis of effects in understudied populations, although researchers should use caution when interpreting the magnitudes of these estimates. This result also motivates the development of more advanced statistical techniques that can be used to integrate and debias cross-population MR estimates.

We observed bias at both more and less stringent instrument selection thresholds. However, bias was less pronounced at the more stringent threshold. We hypothesize that a more stringent threshold increases the proportion of instruments that are causal variants, partially resolving discrepancies due to differences in LD. This observation is supported by recent literature proposing the use of cross-ancestry fine-mapping ^34;35^ to identify causal variants. However, we expect that even perfect causal variant identification will not eliminate bias due to population-mismatch because it does not address effect differences due to gene-environment interactions, and LD differences may still play a role when multiple causal variants are in LD in at least one population.

One limitation of our study results from the disparity in the abundance and quality of GWAS across populations. While we were always able to find at least one European ancestry study for each exposure and outcome trait, this was not true for other populations. For example, there is no available study of Vitamin D, multiple sclerosis, or Alzheimer’s disease in an East Asian population in either OpenGWAS or the GWAS catalog. Summary statistics were even more limited in non-European, non-East Asian populations, including South Asian and African populations. Additionally European ancestry studies have sample sizes between 300,000 and 600,000, while the best or only available studies for South Asian and African populations were typically less than 10,000. As a result, our estimates of population mismatch-induced shrinkage in effect estimates is much less precise for these populations. Data availability for non-European ancestries also limited the power of our replication analysis in Section 4.3. However, even with improved parity in study availability and sample size between populations, we do not expect this limitation to affect the qualitative conclusion that mismatch typically leads to bias towards the null in effect estimates.

To isolate the impact of population mismatch on MR, we compared estimates that differed only by the choice of exposure study. However, in addition to sample population, it is possible that factors related to sample size differences may contribute to differences between MR estimates. We controlled for one of these factors, differential weak-instrument bias, by using MR methods that are robust or immune to this bias (GRAPPLE and MRBEE, respectively). Harbord et al. ^36^ also demonstrate that MR estimates have both asymptotic and small-sample bias when estimating causal odds ratios. However, the shrinkage we observed in our empirical survey was far greater than the bias expected by Harbord et al. ^36^ in estimating the causal odds ratio in typical MR scenarios (around 10% for sample sizes above 2,000). Moreover, we observed shrinkage in population-mismatched MR estimates for nearly all population pairs, regardless of whether the mismatching exposure had a smaller or larger sample size than the matching exposure. This provides strong evidence that sample size differences are not responsible for these results, although sample size differences may affect the exact magnitude of our overall and population pair-specific shrinkage coefficients.

We found a significant, negative association between population-pair specific shrinkage coefficient estimates and *F*_*st*_ distance. However, there is heterogeneity in the degree of shrinkage across trait pairs. We also found that shrinkage due to population mismatch is not necessarily symmetric when exposure and outcome populations are swapped. Differences in the amount of shrinkage observed across trait pairs may be due both to differences in LD discrepancies around instruments and to differences in gene-environment interactions across traits. In this analysis, we have not characterized the relative contributions of these two factors.

## Data Availability

All data produced in the present study are available upon reasonable request to the authors.

https://gwas.mrcieu.ac.uk/

https://www.ebi.ac.uk/gwas/

https://www.finngen.fi/en/access_results

## 6 Acknowledgements

This research was supported by National Human Genome Research Institute (NHGRI) R01 HG013104 and T32-HG000040. We want to acknowledge the participants and investigators of the FinnGen study for access to FinnGen summary data. We thank the work of the GWAS Catalog team and MRC Integrative Epidemiology Unit for making full summary statistics for GWAS studies publicly available through the GWAS Catalog and OpenGWAS projects, respectively. We thank Dr. Michael Boehnke, Dr. Laura Scott, Dr. Sebastian Zoellner, Dr. Hyun Min Kang and other members of the Center of Statistical Genetics at the University of Michigan for their helpful discussions and comments on the manuscript.

## 8 Data and code availability

All GWAS summary statistics used in our MR study are publicly available with links given in Supplemental Data 2 and Supplemental Data 3. Ancestry-specific reference panels from 1000 Genomes used for LD clumping available at http://fileserve.mrcieu.ac.uk/ld/1kg.v3.tgz. Code for reproducing all analysis results is available at https://github.com/lijackk/mismatch-and-2smr.

---

**Table 1.**
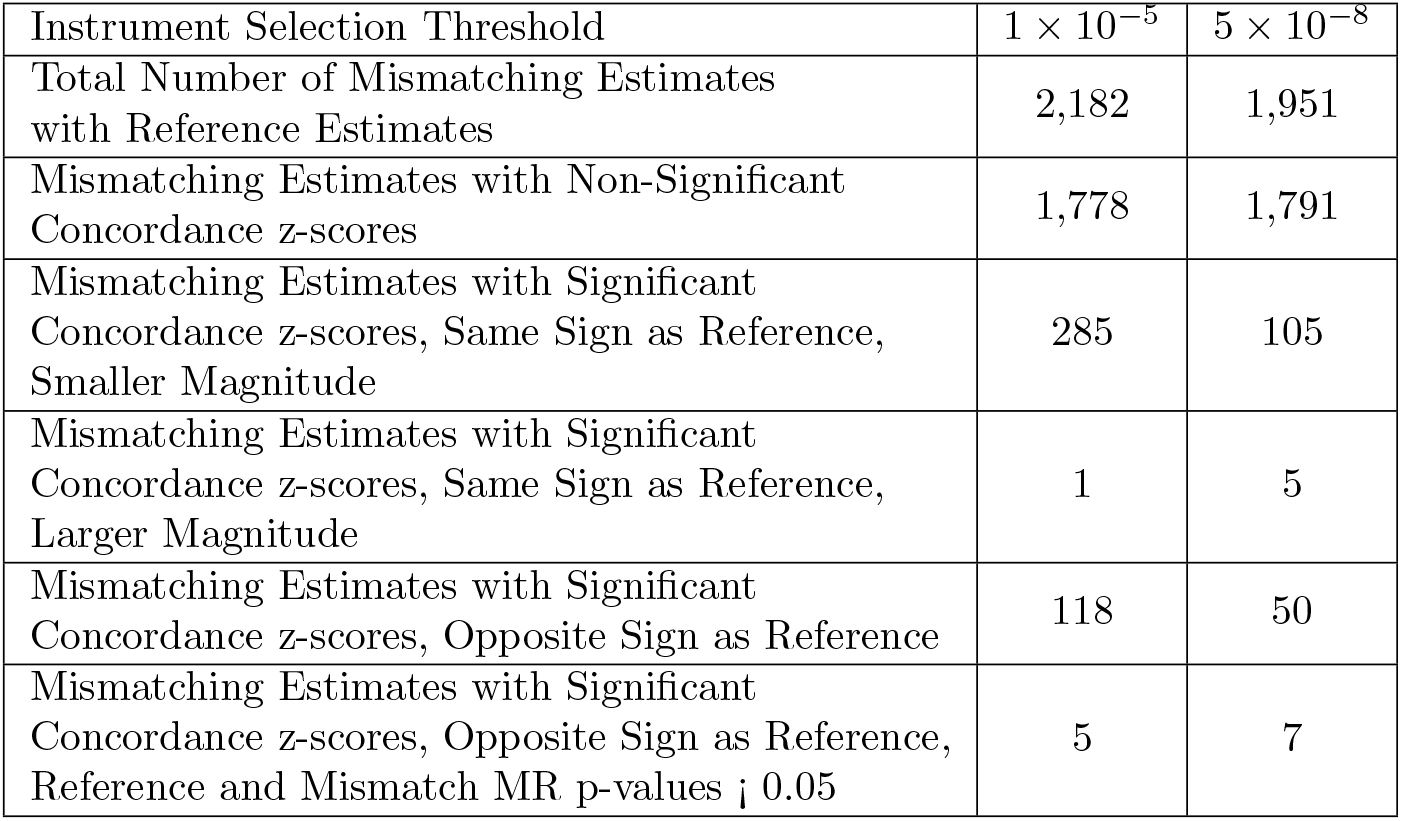
A summary of how MR estimates with mismatching exposures compared with their corresponding reference MR estimates at two different instrument selection thresholds. All estimates were computed using GRAPPLE.

**Table 2.**
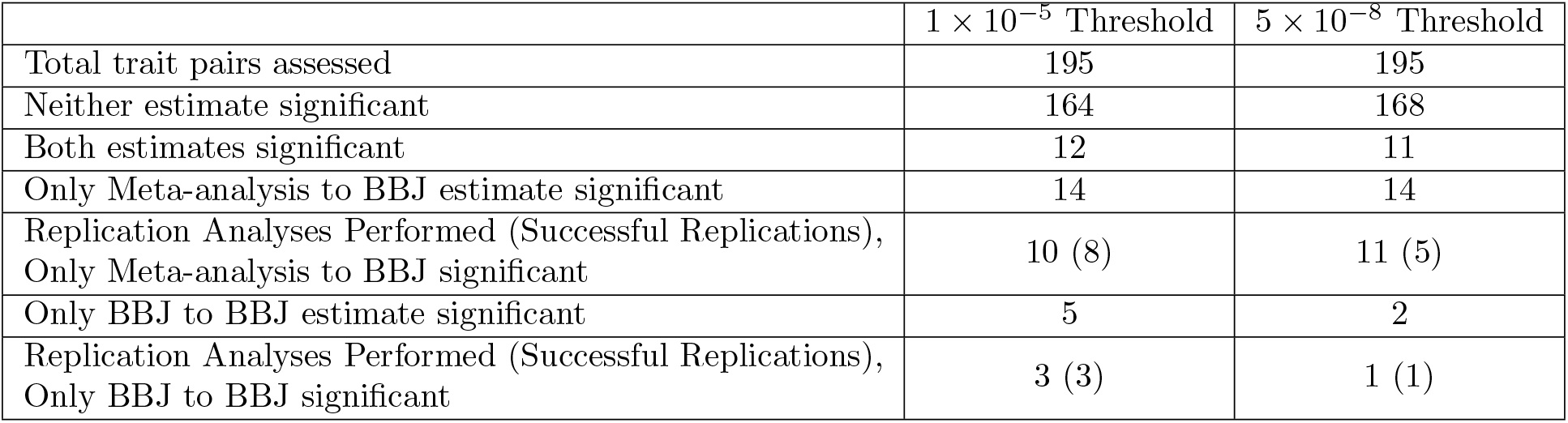
Counts of significant and non-significant causal estimates with outcome studies from Biobank Japan (BBJ) and exposure studies from either BBJ or a meta analysis of BBJ and UK Biobank, as well as replication analysis results for trait pairs that were significant for only one choice of exposure. Significance of MR estimates is based on a Bonferroni-corrected threshold of 0.05/195.

| | $1 \times 10^{-5}$ Threshold | $5 \times 10^{-8}$ Threshold |
| --- | --- | --- |
| Total trait pairs assessed | 195 | 195 |
| Neither estimate significant | 164 | 168 |
| Both estimates significant | 12 | 11 |
| Only Meta-analysis to BBJ estimate significant | 14 | 14 |
| Replication Analyses Performed (Successful Replications),<br>Only Meta-analysis to BBJ significant | 10 (8) | 11 (5) |
| Only BBJ to BBJ estimate significant | 5 | 2 |
| Replication Analyses Performed (Successful Replications),<br>Only BBJ to BBJ significant | 3 (3) | 1 (1) |

## References

[1] Philip C Haycock, Stephen Burgess, Kaitlin H Wade, Jack Bowden, Caroline Relton, and George Davey Smith. Best (but oft-forgotten) practices: the design, analysis, and interpretation of mendelian randomization studies. American Journal of Clinical Nutrition, 103(4): 965–78, 2016.

[2] Maria Cerezo, Elliot Sollis, Yue Ji, Elizabeth Lewis, Ala Abid, Karatug Ozan Bircan, Peggy Hall, James Hayhurst, Sajo John, Abayomi Mosaku, Santhi Ramachandran, Amy Foreman, Arwa Ibrahim, James McLaughlin, Zoë Pendlington, Ray Stefancsik, Samuel A Lambert, Aoife McMahon, Joannella Morales, Thomas Keane, Michael Inouye, Helen Parkinson, and Laura W Harris. The nhgri-ebi gwas catalog: standards for reusability, sustainability and diversity. Nucleic Acids Research, 53(D1):D998–D1005, 2025.

[3] Matthew S. Lyon, Shea J. Andrews, Ben Elsworth, Tom R. Gaunt, Gibran Hemani, and Edoardo Marcora. The variant call format provides efficient and robust storage of gwas summary statistics. Genome Biology, 22(32), 2021.

[4] Stephen Burgess, Neil M. Davies, and Simon G. Thompson. Bias due to participant overlap in two-sample mendelian randomization. Genetic Epidemiology, 40(7): 597–608, 2016.

[5] Jingshu Wang, Qingyuan Zhao, Jack Bowden, Gibran Hemani, George Davey Smith, Dylan S. Small, and Nancy R. Zhang. Causal inference for heritable phenotypic risk factors using heterogeneous genetic instruments. PLoS Genetics, 17(6), 2021.

[6] Jean Morrison, Nicholas Knoblauch, Joseph Marcus, Matthew Stephens, and Xin He. Mendelian randomization accounting for correlated and uncorrelated pleiotropic effects using genome-wide summary statistics. Nature Genetics, 52 (7):740–747, 2020.

[7] Noah Lorincz-Comi, Yihe Yang, Gen Li, and Xiaofeng Zhu. Mrbee: A bias-corrected multivariable mendelian randomization method. HGG Advances, 5(3), 2024.

[8] Stephen Burgess, Robert A. Scott, Nicholas J. Timpson, George Davey Smith, Simon G. Thompson, and EPIC-InterAct Consortium. Using published data in mendelian randomization: a blueprint for efficient identification of causal risk factors. European Journal of Epidemiology, 30(7): 543–552, 2015.

[9] Kevin J. Galinsky, Yakir A. Reshef, Hilary K. Finucane, Po-Ru Loh, Noah Zaitlen, Nick J. Patterson, Brielin C. Brown, and Alkes L. Price. Estimating cross-population genetic correlations of causal effect sizes. Genetic Epidemiology, 43(2): 180–188, 2018.

[10] Ying Wang, Jing Guo, Guiyan Ni, Jian Yang, Peter M. Visscher, and Loic Yengo. Theoretical and empirical quantification of the accuracy of polygenic scores in ancestry divergent populations. Nature Communications, 11:3865, July 2020. ISSN 2041-1723. doi: 10.1038/s41467-020-17719-y. URL https://pmc.ncbi.nlm.nih.gov/articles/PMC7395791/.

[11] Qingyuan Zhao, Jingshu Wang, Wes Spiller, Jack Bowden, and Dylan S. Small. Two-Sample Instrumental Variable Analyses Using Heterogeneous Samples. Statistical Science, 34(2):317–333, May 2019. ISSN 0883-4237, 2168-8745. doi: 10.1214/18-STS692. URL https://projecteuclid.org/journals/statistical-science/volume-34/issue-2/Two-Sample-Instrumental-Variable-Analyses-Using-Heterogeneous-Samples/10.1214/18-STS692.full. Publisher: Institute of Mathematical Statistics.

[12] Benjamin Woolf, Amy Mason, Loukas Zagkos, Hannah Sallis, Marcus R. Munafo, and Dipender Gill. MRSamePopTest: introducing a simple falsification test for the two-sample mendelian randomisation ‘same population’ assumption. BMC research notes, 17(1):27, January 2024. ISSN 1756-0500. doi: 10.1186/s13104-024-06684-0.

[13] Veronika W Skrivankova, Rebecca C Richmond, Benjamin A R Woolf, James Yarmolinsky, Neil M Davies, Sonja A Swanson, Tyler J VanderWeele, Julian P T Higgins, Nicholas J Timpson, Niki Dimou, Claudia Langenberg, Robert M Golub, Elizabeth W Loder, Valentina Gallo, Anne Tybjaerg-Hansen, George Davey Smith, Matthias Egger, and J Brent Richards. Strengthening the reporting of observational studies in epidemiology using mendelian randomization (strobe-mr) statement. JAMA, 326(16): 1614–1621, 2021.

[14] Huiling Zhao, Humaria Rasheed, Therese Haugdahl Nøst, Yoonsu Cho, Yi Liu, Laxmi Bhatta, Arjun Bhattacharya, Global Biobank Meta analysis Initiative, Gibran Hemani, George Davey Smith, Ben Michael Brumpton, Wei Zhou, Benjamin M Neale, Tom R Gaunt, and Jie Zheng. Proteome-wide mendelian randomization in global biobank meta-analysis reveals multi-ancestry drug targets for common diseases. Cell Genomics, 2(11), 2022.

[15] Elsa Valdes-Marquez, Sarah Parish, Robert Clarke, Traiani Stari, Bradford B Worrall, METASTROKE Consortium of the ISGC, and Jemma C Hopewell. Relative effects of ldl-c on ischemic stroke and coronary disease. Neurology, 92 (11):1176–1187, 2019.

[16] Pengsheng Li, Haiyan Wang, Lan Guo, Xiaoyan Gou, Gengdong Chen, Dongxin Lin, Dazhi Fan, Xiaoling Guo, and Zhengping Liu. Association between gut microbiota and preeclampsia-eclampsia: a two-sample mendelian randomization study. BMC Medicine, 20(1): 443, 2022.

[17] Yiwen Long, Lanhua Tang, Yangying Zhou, Shushan Zhao, and Hong Zhu. Causal relationship between gut microbiota and cancers: a two-sample mendelian randomisation study. BMC Medicine, 21(1): 66, 2023.

[18] Fernando Pires Hartwig, Maria Carolina Borges, Bernardo Lessa Horta, Jack Bowden, and George Davey Smith. Inflammatory biomarkers and risk of schizophrenia: A 2-sample mendelian randomization study. JAMA Psychiatry, 74 (12):1226–1233, 2017.

[19] Holmes MV, Asselbergs FW, Palmer TM, Drenos F, Lanktree MB, Nelson CP, Dale CE, Padmanabhan S, Finan C, Swerdlow DI, Tragante V, van Iperen EP, Sivapalaratnam S, Shah S, Elbers CC, Shah T, Engmann J, Giambartolomei C, White J, Zabaneh D, Sofat R, McLachlan S.; UCLEB consortium.; Doevendans PA, Balmforth AJ, Hall AS, North KE, Almoguera B, Hoogeveen RC, Cushman M, Fornage M, Patel SR, Redline S, Siscovick DS, Tsai MY, Karczewski KJ, Hofker MH, Verschuren WM, Bots ML, van der Schouw YT, Melander O, Dominiczak AF, Morris R, Ben-Shlomo Y, Price J, Kumari M, Baumert J, Peters A, Thorand B, Koenig W, Gaunt TR, Humphries SE, Clarke R, Watkins H, Farrall M, Wilson JG, Rich SS, de Bakker PI, Lange LA, Davey Smith G, Reiner AP, Talmud PJ, Kivimäki M, Lawlor DA, Dudbridge F, Samani NJ, Keating BJ, Hingorani AD, and Casas JP. Mendelian randomization of blood lipids for coronary heart disease. European Heart Journal, 36(9): 539–550, 2015.

[20] Yi Ding, Kangcheng Hou, Ziqi Xu, Aditya Pimplaskar, Ella Petter, Kristin Boulier, Florian Privé, Bjarni J. Vilhjálmsson, Loes M. Olde Loohuis, and Bogdan Pasaniuc. Polygenic scoring accuracy varies across the genetic ancestry continuum. Nature, 618(7966):774–781, June 2023. ISSN 1476-4687. doi: 10.1038/s41586-023-06079-4. URL https://www.nature.com/articles/s41586-023-06079-4. Publisher: Nature Publishing Group.

[21] Eleanor Sanderson, M. Mario Glymour, Michael Holmes, Hyunseung Kang, Jean Morrison, Marcus Munafo, Tom Palmer, C. Mary Schooling, Chris Wallace, Qingyuan Zhao, and George Davey Smith. Mendelian randomization. Nature Review Methods Primers, 2(6), 2022.

[22] Fernando Pires Hartwig, Linbo Wang, George Davey Smith, and Neil Martin Davies. Average Causal Effect Estimation Via Instrumental Variables: the No Simultaneous Heterogeneity Assumption. Epidemiology, 34(3):325, May 2023. ISSN 1044-3983. doi: 10.1097/EDE.0000000000001596. URL https://journals.lww.com/epidem/abstract/2023/05000/average_causal_effect_estimation_via_instrumental.4.aspx.

[23] Stephen Burgess and Jeremy A. Labrecque. Mendelian randomization with a binary exposure variable: interpretation and presentation of causal estimates. European Journal of Epidemiology, 33(10): 947–952, 2018. ISSN 0393-2990. doi: 10.1007/s10654-018-0424-6. URL https://pmc.ncbi.nlm.nih.gov/articles/PMC6153517/.

[24] Stephen Burgess, Adam Butterworth, and Simon G Thompson. Mendelian randomization analysis with multiple genetic variants using summarized data. Genetic Epidemiology, 37(7): 658–665, 2013.

[25] Jack Bowden, George Davey Smith, Philip Haycock, and Stephen Burgess. Consistent estimation in mendelian randomization with some invalid instruments using a weighted median estimator. Genetic Epidemiology, 40(4): 304–314, 2016.

[26] Xiang Zhu and Matthew Stephens. Bayesian Large-Scale Multiple Regression With Summary Statistics From Genome-Wide Association Studies. The annals of applied statistics, 11(3): 1561–1592, 2017. ISSN 1932-6157. doi: 10.1214/17-aoas1046. URL https://pmc.ncbi.nlm.nih.gov/articles/PMC5796536/.

[27] Rui Xiao and Michael Boehnke. Quantifying and correcting for the winner’s curse in genetic association studies. Genetic Epidemiology, 33(5): 453–462, 2009.

[28] Cameron Palmer and Itsik Pe’er. Statistical correction of the winner’s curse explains replication variability in quantitative trait genome-wide association studies. PLoS Genetics, 13(7):e1006916, 2017.

[29] Mitja I. Kurki, Juha Karjalainen, Priit Palta, Timo P. Sipilä, Kati Kristiansson, Kati M. Donner, Mary P. Reeve, Hannele Laivuori, Mervi Aavikko, Mari A. Kaunisto, Anu Loukola, Elisa Lahtela, Hannele Mattsson, Päivi Laiho, Pietro Della Briotta Parolo, Arto A. Lehisto, Masahiro Kanai, Nina Mars, Joel Rämö, Tuomo Kiiskinen, Henrike O. Heyne, Kumar Veerapen, Sina Ruëger, Susanna Lemmelä, FinnGen, …, and Aarno Palotie. Finngen provides genetic insights from a well-phenotyped isolated population. Nature, 613: 508–518, 2023.

[30] J.R. Cook and L.A. Stefanski. Simulation-extrapolation estimation in parametric measurement error models. Journal of the American Statistical Association, 89(428): 1314–1328, 1994.

[31] Eric M Scott, Anason Halees, Yuval Itan, Emily G Spencer, Yupeng He, Mostafa Abdellateef Azab, Stacey B Gabriel, Aziz Belkadi, Bertrand Boisson, Laurent Abel, Andrew G Clark, Greater Middle East Variome Consortium, Fowzan S Alkuraya, Jean-Laurent Casanova, and Joseph G Gleeson. Characterization of greater middle eastern genetic variation for enhanced disease gene discovery. Nature Genetics, 48(9): 1071–1076, 2016.

[32] Saori Sakaue, Masahiro Kanai, Yosuke Tanigawa, Juha Karjalainen, Mitja Kurki, Seizo Koshiba, Akira Narita, Takahiro Konuma, Kenichi Yamamoto, Masato Akiyama, Kazuyoshi Ishigaki, Akari Suzuki, Ken Suzuki, Wataru Obara, Ken Yamaji, Kazuhisa Takahashi, Satoshi Asai, Yasuo Takahashi, Takao Suzuki, Nobuaki Shinozaki, Hiroki Yamaguchi, Shiro Minami, Shigeo Murayama, Kozo Yoshimori, Satoshi Nagayama, Daisuke Obata, Masahiko Higashiyama, Akihide Masumoto, Yukihiro Koretsune, FinnGen Kaoru Ito, Chikashi Terao, Toshimasa Yamauchi, Issei Komuro, Takashi Kadowaki, Gen Tamiya, Masayuki Yamamoto, Yusuke Nakamura, Michiaki Kubo, Yoshinori Murakami, Kazuhiko Yamamoto, Yoichiro Kamatani, Aarno Palotie, Manuel A Rivas, Mark J Daly, Koichi Matsuda, and Yukinori Okada. A cross-population atlas of genetic associations for 220 human phenotypes. Nature Genetics, 53(10): 1415–1424, 2021.

[33] Kazuyoshi Ishigaki, Saori Sakaue, Chikashi Terao, Yang Luo, Kyuto Sonehara, Kensuke Yamaguchi, Tiffany Amariuta, Chun Lai Too, Vincent A. Laufer, Ian C. Scott, Sebastien Viatte, Meiko Takahashi, Koichiro Ohmura, Akira Murasawa, Motomu Hashimoto, Hiromu Ito, Mohammed Hammoudeh, Samar Al Emadi, Basel K. Masri, Hussein Halabi, Humeira Badsha, Imad W. Uthman, Xin Wu, Li Lin, Ting Li, Darren Plant, Anne Barton, Gisela Orozco, Suzanne M. M. Verstappen, John Bowes, Alexander J. MacGregor, Suguru Honda, Masaru Koido, Kohei Tomizuka, Yoichiro Kamatani, Hiroaki Tanaka, Eiichi Tanaka, Akari Suzuki, Yuichi Maeda, Kenichi Yamamoto, Satoru Miyawaki, Gang Xie, Jinyi Zhang, Christopher I. Amos, Edward Keystone, Gertjan Wolbink, Irene van der Horst-Bruinsma, Jing Cui, Katherine P. Liao, Robert J. Carroll, Hye-Soon Lee, So-Young Bang, Katherine A. Siminovitch, Niek de Vries, Lars Alfredsson, Solbritt Rantapää-Dahlqvist, Elizabeth W. Karlson, Sang-Cheol Bae, Robert P. Kimberly, Jeffrey C. Edberg, Xavier Mariette, Tom Huizinga, Philippe Dieudé, Matthias Schneider, Martin Kerick, Joshua C. Denny, Koichi Matsuda, Keitaro Matsuo, Tsuneyo Mimori, Fumihiko Matsuda, Keishi Fujio, Yoshiya Tanaka, Atsushi Kumanogoh, Matthew Traylor, Cathryn M. Lewis, Stephen Eyre, Huji Xu, Richa Saxena, Thurayya Arayssi, Yuta Kochi, Katsunori Ikari, Masayoshi Harigai, Peter K. Gregersen, Kazuhiko Yamamoto, S. Louis Bridges, Leonid Padyukov, Javier Martin, Lars Klareskog, Yukinori Okada, and Soumya Raychaudhuri. Multi-ancestry genome-wide association analyses identify novel genetic mechanisms in rheumatoid arthritis. Nature Genetics, 54(11):1640–1651, November 2022. ISSN 1546-1718. doi: 10.1038/s41588-022-01213-w. URL https://www.nature.com/articles/s41588-022-01213-w. Publisher: Nature Publishing Group.

[34] Gibran Hemani, Yoonsu Cho, Amanda Chong, Tom Palmer, Amy Mason, John Ferguson, David Evans, and George Davey Smith. Jointly modelling multiple ancestral populations using gwas summary data improves causal inference. biorXiv [PREPRINT], 2025.

[35] Yihe Yang and Xiaofeng Zhu. Improving causal effect estimation in multi-ancestry multivariable mendelian randomization with transfer learning. bioRxiv, 2025. doi: 10.1101/2025.07.11.664423.

[36] Roger M. Harbord, Vanessa Didelez, Tom M. Palmer, Sha Meng, Jonathan A. C. Sterne, and Nuala A. Sheehan. Severity of bias of a simple estimator of the causal odds ratio in Mendelian randomization studies. Statistics in Medicine, 32(7): 1246–1258, March 2013. ISSN 1097-0258. doi: 10.1002/sim.5659.

