## Supplemental Note, Tables and Figures for "Mind the gap: characterizing bias due to population mismatch in two-sample Mendelian randomization"

#### 1 Supplemental Methods

##### 1.1 Nonlinearity and homogeneity assumptions

In Methods Section "Statistical Setting and Assumptions", we discuss MR estimation when exposure, $X$ and outcome, $Y$ are related by a linear structural equation model $Y_{i,t} = \gamma_t X_{i,t} + v_{i,p}$ for individual $i$ in the target population. In this case, the MR estimate is interpretable as an estimate of $\gamma_t$. If $X$ and $Y$ are related by a nonlinear structural equation, $Y_{i,t} = g_t(X_{i,t}, v_{i,p})$, the quantity estimated by MR can be interpreted as a weighted average of partial derivatives of $g_t$ with respect to $X$ at individual-specific values of the exposure $X$[1;2]. If $g_t$ is monotonic for all $v_{i,t}$, then the MR estimate provides a valid test of sign and significance. The weights in the weighted average depend on the effect size of the instrumental variables used. This means that, theoretically, two MR estimates based on the same outcome data but different sets of instruments could have different estimands, even if variant-exposure associations transfer perfectly between exposure and outcome populations. However, MR studies typically use a large number of variants with small effects, so we expect differences in instrument effects sizes to have a very small impact on differences in the MR estimate across analyses.

Section "Statistical Setting and Assumptions" also describes several variants of the the fourth MR assumption. Often MR literature assumes homogeneity of the $G - X$ association. For example, this variation of the fourth assumption is satisfied if $G_1, \dots, G_J$ and $X$ are related by the homogeneous, linear structural equation model $X_{i,t} = \sum_{j=1}^{J} \beta_{j,t} G_{j,i,t} + u_{i,t}$. However, homogeneity and linearity are much stronger conditions than are needed to justify MR estimation. One alternative is monotonicity, which requires that the linear association between $G_j$ and $X$ is either non-negative or non-positive for all individuals[3;4;5]. If the association between $G_j$ and $X$ is strictly positive or strictly negative for all individuals and all variants, and either $X$ is binary or the $X - Y$ relationship is linear for all individuals, then IVW regression and related MR estimators estimate, the average treatment effect in the target population. However, if the association between $G_j$ and $X$ is zero for some individuals and some instruments, then each Wald ratio, $\hat{\beta}_{Y,j,t}/\hat{\beta}_{X,j,t}$, estimates the local average treatment effect (LATE), or the average treatment effect among individuals with non-zero association between $G_j$ and $X$ (compliers). In MR studies, we make use of many instruments. As a result the final estimate estimates a weighted average of LATEs corresponding to the complier set for each instrument.

A third variation of the fourth assumption described by Hartwig et al.[5] is the no simultaneous heterogeneity (NOSH) assumption, which requires heterogeneity in the causal effect of $X$ on $Y$ to be uncorrelated with both the instrument $G_j$ and any heterogeneity in the $G_j - X$ association. Hartwig et al.[5] demonstrate that Wald ratios for variants that satisfy this assumption are consistent for the average treatment effect. The NOSH assumption is plausible in MR settings, where there is no reason to expect simultaneous heterogeneity in variant-exposure and exposure-outcome relationships. Under the NOSH assumption, we would not expect the estimand of MR estimates made with two different sets of variants to differ, even if the instruments chosen had different complier sets.

##### 1.2 Survivorship bias in Mendelian randomization

In Methods Section "Typical IV selection practices induce shrinkage in MR with mismatching populations", we claim that selecting instruments based on the exposure population results in variants with larger magnitude exposure associations in the exposure population. In other words, if $S = 1$ indicates the event that $G_j$ is selected as an instrument using data from the exposure population, we claim that $E[\beta^2_{X,j,e}|S = 1] > E[\beta^2_{X,j,t}|S = 1]$.

To improve readability, we will prove this result dropping the subscript $j$. We first assume that,

$$\beta_{X,e} = \beta + \mu_e \qquad (1)$$
$$\beta_{X,t} = \beta + \mu_t, \qquad (2)$$

where $\mu_e$, $\mu_t$, $\beta$ are mutually independent, $\mu_t$ and $\mu_e$ are mean zero and symmetrically distributed around zero, and $\beta$ is mean zero, symmetric, and unimodal. In this parameterization, $\beta$ is the average average association between the exposure and target population and $\mu_e$ and $\mu_t$ are population-specific deviations from this central value. We further assume that $\mu_t$ and $\mu_e$

have equal variance, denoted as $V_\mu$. We also assume that a variant is selected if the $z$-score for an estimate of $\beta_{X,e}$, $\hat{\beta}_{X,e}$ exceeds a positive threshold in magnitude,

$$\frac{|\hat{\beta}_{X,e}|}{\hat{s}_e} = \frac{|\beta + \mu_e + \epsilon|}{\hat{s}_e} > T', \tag{3}$$

where $\epsilon$ is a mean zero, symmetrically distributed, unimodal error term independent of $\beta, \mu_t, \mu_e$, and $\hat{s}_e$ is an estimate of the variance of $\epsilon$ that is independent of $\beta$, $\mu_e$, and $\epsilon$. Since $\hat{s}_e$ is independent of other conditions, we can simplify the selection condition to

$$|\hat{\beta}_{X,e}| = |\beta + \mu_e + \epsilon| > T, \tag{4}$$

where $T = \hat{s}_e T'$.

To prove our claim, we will show that $E[\beta^2_{X,e} - \beta^2_{X,t}|S = 1] > 0$. We first decompose this expression as

$$E[\beta^2_{X,e} - \beta^2_{X,t}|S = 1] = E[\beta^2 + 2\mu_e\beta + \mu_e^2 - \beta^2 - 2\mu_t\beta - \mu_t^2|S = 1] = E[2\mu_e\beta - 2\mu_t\beta + \mu_e^2 - \mu_t^2|S = 1]. \tag{5}$$

Because $\mu_t$ is independent of the selection event,

$$E[\mu_t^2|S = 1] = E[\mu_t^2] = V_\mu \tag{6}$$

$$E[\beta\mu_t|S = 1] = E_\beta[\beta E_{\mu_t}[\mu_t|S = 1, \beta]|S = 1] = E_\beta[\beta \cdot 0|S = 1] = 0. \tag{7}$$

Next, we prove that $E[\mu_e^2|S = 1] \geq E[\mu_e^2] = V_\mu$. First, note that $E[\mu_e^2|S = 1] = \frac{1}{P[S=1]}E[\mu_e^2 1_{S=1}]$ and that $E[\mu_e^2 1_{S=1}] = E_{\mu_e}[\mu_e P[S = 1|\mu_e]]$. Since $B = \beta + \epsilon$ is symmetric about 0, for any $m > 0$,

$$\begin{aligned} P[S = 1|\mu_e = m] =& 1 - P[B \in (-T - m, T - m)] \\ =& 1 - P[B \in (-T + m, T + m)] = P[S = 1|\mu_e = -m], \end{aligned} \tag{8, 9}$$

so $P[S = 1|\mu_e = m]$ is symmetric as a function of $m$. If $B$ is also unimodal, then for any $m > m' > 0$,

$$\begin{aligned} P[S =& 1|\mu_e = m] = 1 - P[B \in (-T - m, T - m)] && (10)\\ =& 1 - \{P[B \in (-T - m, -T - m')] + P[B \in (-T - m', T - m')] - P[B \in (T - m, T - m')]\} && (11)\\ =& P[S = 1|\mu_e = m'] - \{P[B \in (T + m', T + m)] - P[B \in (T - m, T - m')]\} && (12)\\ \geq& P[S = 1|\mu_e = m']. && (13) \end{aligned}$$

This shows that $P[S = 1|\mu_e]$ can be written as a monotonically increasing function of $|\mu_e|$. Since $\mu_e^2$ is also a monotonically increasing function of $|\mu_e|$, by the Chebyshev integral inequality for weighted measures[6],

$$E[\mu_e^2|S = 1] = \frac{E[\mu_e^2 P[S = 1|\mu_e]]}{P[S = 1]} \geq \frac{E[\mu_e^2]E[P[S = 1|\mu_e]]}{P[S = 1]} = V_\mu. \tag{14}$$

Finally, we show that $E[\beta\mu_e|S = 1] > 0$. Using the iterated expectation formula

$$E[\beta\mu_e|S = 1] = E_\beta[\beta E[\mu_e|S = 1, \beta]] \tag{15}$$

We will show that $sign(E[\mu_e|S = 1, \beta]) = sign(\beta)$, meaning that $\beta E[\mu_e|S = 1, \beta] > 0$ for all values of $\beta$. Let $f_{\mu_e,\epsilon|S=1,\beta}$ be the joint conditional density of $\mu_e$ and $\epsilon$ given $S = 1$ and $\beta$. Using Bayes rule and the joint independence of $\mu_e$, $\epsilon$, and $\beta$,

$$f_{\mu_e,\epsilon|S=1,\beta}(m, y) = \frac{P[S = 1|\mu_e = m, \epsilon = y, \beta]f_{\mu_e,\epsilon|\beta}(m, y|\beta)}{P[S = 1|\beta]} = \frac{1_{|\beta+m+y|>T}f_{\mu_e}(m)f_\epsilon(y)}{P[S = 1|\beta]}, \tag{16}$$

where $f_{\mu_e}$ and $f_\epsilon$ are the unconditional marginal densities of $\mu_e$ and $\epsilon$. Therefore,

$$\begin{aligned} E[\mu_e|S = 1, \beta] =& \int_{-\infty}^{\infty}\int_{-\infty}^{\infty} m f_{\mu_e,\epsilon|S=1,\beta}(m, y)dy\ dm && (17)\\ =& \frac{1}{P[S = 1|\beta]}\int_{-\infty}^{\infty} m f_{\mu_e}(m)\left(\int_{-\infty}^{\infty} 1_{|\beta+m+y|>T}f_\epsilon(y)dy\right) dm && (18)\\ =& \frac{1}{P[S = 1|\beta]}\int_{-\infty}^{\infty} m\left(1 - P[\epsilon \in (-T - \beta - m, T - \beta - m)]\right) f_{\mu_e}(m)dm. && (19) \end{aligned}$$

Let $g(m,\beta) = P[\epsilon \in (-T-\beta-m, -T-\beta-m)]$. Since the distribution of $\epsilon$ is symmetric, $g(m,\beta)$ is symmetric about $m = -\beta$. Furthermore, because the distribution of $\epsilon$ is unimodal, $g(m,\beta) > g(m',\beta)$ if $|\beta+m| > |\beta+m'|$. Using symmetry of the distribution of $\mu_e$,

$$E[\mu_e|S=1,\beta] = \frac{1}{P[S=1|\beta]}\int_{-\infty}^{\infty} m f_{\mu_e}(m) g(m,\beta) dm \tag{20}$$

$$= \frac{1}{P[S=1|\beta]}\left\{\int_{-\infty}^{0} m f_{\mu_e}(m) g(m,\beta) dm + \int_{0}^{\infty} m f_{\mu_e}(m) g(m,\beta) dm\right\} \tag{21}$$

$$= \frac{1}{P[S=1|\beta]}\int_{0}^{\infty} m f_{\mu_e}(m)\left(g(m,\beta) - g(-m,\beta)\right) dm. \tag{22}$$

If $\beta > 0$, then $|\beta+m| > |\beta-m|$ for all positive $m$, so $E[\mu_e|S=1,\beta] > 0$. Conversely, if $\beta < 0$, then $|\beta+m| < |\beta-m|$ for all positive $m$, so $E[\mu_e|S=1,\beta] < 0$. Therefore, $sign(E[\beta\mu_e|S=1]) = sign(\beta)$, completing the proof.

Combining these results shows that

$$E[\beta_{X,j,e}^2 - \beta_{X,j,t}^2|S=1] = 2E[\beta\mu_e|S=1] + E[\mu_e^2 - \mu_t^2|S=1], \tag{23}$$

where $E[\beta\mu_e|S=1]$ is positive and $E[\mu_e^2 - \mu_t^2|S=1]$ is non-negative.

This argument requires some mild distributional assumptions about $\beta_{X,j,e}$ and $\beta_{X,j,t}$. We assumed that population-specific deviations from the average effect across populations are mean zero, symmetrically distributed, and equal variance across populations. This is a common assumption made in meta-analyses of GWAS. We also assumed that the average effects, $\beta$ are mean zero, symmetrically distributed, and additionally assumed this distribution to be unimodal. These assumptions are mild and common to almost all previous work modeling the distribution of causal genetic effects[7;8;9]. Finally, $\epsilon$, representing a random noise term in GWAS estimation, is expected to be asymptotically normal, consistent with our assumptions that its distribution is mean zero, symmetric, and unimodal.

When populations are mismatched and instruments are selected based on $\hat{\beta}_{X,j,e}$, both winner's curse and survivorship bias contribute to the average deflation of MR estimates through similar but separate mechanisms. Winner's curse bias occurs when variants are selected as instruments using the same data used to compute the MR estimate. In this case, $\hat{\beta}_{X,j,e}$ will, on average, overestimate $\beta_{X,j,e}$. However, even if a separate sample from the exposure population is used to select instruments, $\beta_{X,j,e}$ will be, on average, larger in magnitude than $\beta_{X,j,t}$ due to survivorship bias.

### 2 Mendelian randomization analysis

For each MR analysis, we identified genetic variants contained in both the exposure and outcome GWAS. We then LD clumped these overlapping variants, prioritizing variants based on their exposure study association p-value using an $r^2$ threshold of 0.01 and distance threshold of 500 kb. We selected the lead variant in each clump to be an instrumental variable if its p-value was less than either $10^{-5}$ or $5 \times 10^{-8}$. For each exposure, we used reference panels from 1000 Genomes from one of the five superpopulations available (AFR, AMR, EAS, EUR, SAS). In the event that an exposure GWAS contains samples from multiple ancestries, we used the reference panel of the superpopulation that made up a majority or plurality of the study cohort (so, for example, a meta-analysis of UK Biobank and Biobank Japan would use a reference panel from EUR). We then merged and harmonized exposure and outcome association estimates for the selected instruments.

To address horizontal pleiotropy in GRAPPLE, we included an overdispersion term, and used a Tukey loss function. We note that MRBEE is also capable of performing pleiotropy detection via removal of invalid instruments[10]. However, including pleiotropy detection meant we were unable to run MR for certain pairs of exposure and outcome studies that we could for GRAPPLE, especially for study pairs with small numbers of instruments ($< 5$). Therefore, the results we present for MRBEE in Table S5 are run without pleiotropy detection, which we accomplished by setting a negative p-value threshold for pleiotropy detection in the `MRBEE.IMRP.UV` function of the MRBEE package[10]. To address sample overlap, we estimated the residual correlation between exposure and outcome effect estimates using the p-value thresholding approach described by Wang et al.[11]. However, we used a threshold of $p > 0.05$ instead of $p > 0.50$ as suggested by Wang et al.[11], as the latter tends to underestimate this shared correlation. We used this matrix as an input for both GRAPPLE and MRBEE. For MR with standard inverse variance weighted (IVW) regression, we used the implementation in the `mr_ivw` function of the TwoSampleMR package[12].

We performed Steiger filtering on selected IVs for each study pair to remove variants more strongly correlated with the outcome than the exposure[13]. Steiger filtering can remove pleiotropic variants and reduce the risk of false positives due to reverse causation. In the Steiger filtering procedure, the proportions of exposure and outcome variance explained by each SNP are estimated and variants are removed if the proportion of outcome variance explained excedes the proportion of exposure variance explained. For continuous traits, the proportion of variance explained by variant $G_j$ can be approximated as

$$R_j^2 \approx \frac{F_j}{F_j - 2 + n},$$

where $n$ is the sample size of the GWAS study, $F_j = (\frac{\hat{\beta}_j}{\hat{se}(\hat{\beta}_j)})^2$, $\hat{\beta}_j$ is the estimated association of $G_j$ with the trait, and $\hat{se}(\hat{\beta}_j)$ is the corresponding estimate of the standard error. For binary traits, we estimated the proportion of variance explained using the `get_r_from_lor()` function in the TwoSampleMR package, which implements an approximation provided by Lee et al. [14]. This function requires the estimated log odds ratio, case and control counts, the effect allele frequency of the variant, and the estimated prevalence of the binary trait in the study population. Prevalences for outcome studies were estimated using external literature, as detailed in Supplemental Data 4. Effect allele frequencies were not provided for four majority European ancestry outcome studies. For these studies, we approximated effect allele frequencies using the EUR superpopulation of 1000 Genomes.

### 3 Estimation with Measurement Error using Simulation Extrapolation (SIMEX)

To estimate the overall and population-pair specific shrinkage coefficients described in the Methods, we performed regression of population-mismatched MR estimates against their corresponding exact-matching reference estimates. To account for noise in the reference estimates, we used simulation extrapolation (SIMEX) [15]. In the SIMEX procedure, we add simulated noise to our real data and recompute the inverse-variance weighted regression coefficient. We perform this procedure for a range of noise level and use the trend in estimates to extrapolate the estimate we would have obtained if the reference estimates had been measured without noise.

Let $\hat{\gamma}_{1,mm}, \ldots, \hat{\gamma}_{J,mm}$ be the population-mismatched estimates used in the analysis. These serve as the outcome in the inverse-variance weighted regression. Let $\hat{\gamma}_{1,r}, \ldots, \hat{\gamma}_{J,r}$ be the corresponding reference (population-matched) estimates, which serve as the covariate. For a grid of values, $\lambda$, between 0 and 5 in intervals of 0.25, we sample new population-mismatched and reference estimates, $\hat{\gamma}_{j,mm,\lambda} \sim N(\hat{\gamma}_{j,mm}, \sigma^2_{j,mm})$ and $\hat{\gamma}_{j,r,\lambda} \sim N(\hat{\gamma}_{j,r}, (1+\lambda)\sigma^2_{j,r})$, where $\sigma_{j,r}$ and $\sigma_{j,mm}$ are the estimated standard errors of $\hat{\gamma}_{j,r}$ and $\hat{\gamma}_{j,mm}$ respectively. For each value of $\lambda$, we compute $\hat{\beta}_\lambda$, the coefficient in the inverse-variance weighted regression. The condition of no measurement error in $\hat{\gamma}_{j,r}$ corresponds to the condition that $\lambda = -1$, which is not observable. We therefore extrapolate this value by fitting a function to approximate $\hat{\beta}_\lambda$ as a function of $\lambda$ and then plugging in $\lambda = -1$. We found that the a linear function fit the data well for our overall shrinkage coefficients (Figure S3), as well as for population pair-specific shrinkage coefficients with many estimate groups available (such as using a UK Biobank exposure for a Biobank Japan target). For population pairs with fewer estimate groups available or less precise regression results, observed data were generally too noisy to rule out a linear extrapolation function. To estimate a standard error for $\hat{\beta}_{-1}$, we repeat this procedure several times resampling the observations that contribute to the regression estimate. The standard error of $\hat{\beta}_{-1}$ is estimated as the standard deviation of the bootstrap estimates.

### 4 Population pair-specific analyses at a more stringent instrument selection threshold

We used SIMEX to estimate population pair-specific shrinkage coefficients using the instrument selection p-value threshold of $5 \times 10^{-8}$ in addition to the more lenient $10^{-5}$ threshold. However, at the stricter threshold, no population pair with an African ancestry GWAS used as the outcome had more than 8 population-mismatched estimates that could be compared to a reference, leaving only 23 population pairs in which SIMEX estimates were computed. The slope of inverse variance weighted regression of estimated shrinkage coefficients vs $F_{st}$ is -2.51, with a standard error of 0.70 ($p = 0.003$). This is similar to the slope of -2.40 obtained when using more lenient threshold of $1 \times 10^{-5}$.

SIMEX-estimated shrinkage coefficients and bootstrap standard errors using both instrument selection thresholds are shown in Supplementary Figure S2. Supplementary Figure Figure S4 shows reference vs population-mismatched estimates for each population pair. At both thresholds, most of these slopes are much lower than 1, aligning with our predictions and the overall trends observed in the main manuscript. In some cases, the SIMEX-estimated shrinkage coefficient is negative, usually when using GWAS sampled from Africans in the Pan-UKB project as the exposure or target population. However, 95% confidence intervals of these coefficients always included zero.

There are some differences in shrinkage coefficients between instrument selection thresholds. For example, the FinnGen exposure - UK Biobank outcome is lower at the more lenient threshold (0.53 for $1 \times 10^{-5}$ vs. 0.65 for $5 \times 10^{-8}$), while the UK Biobank exposure - Biobank Japan outcome estimate is larger at the more lenient threshold (0.44 for $1 \times 10^{-5}$ vs. 0.26 for $5 \times 10^{-8}$).

### 5 Supplemental Tables

| Trait | Matching Outcome | Standard unit | Number of studies |
|---|---|---|---|
| LDL cholesterol | Coronary artery disease | mmol/L | 5 |
| Body Mass Index | Type 2 diabetes | $kg/m^2$ | 6 |
| Hemoglobin A1c | Coronary artery disease | mmol/mol | 4 |
| Systolic Blood Pressure | Ischemic stroke | mmHg | 5 |
| Eosinophil count | Rheumatoid arthritis | $10^9$ cells/L | 4 |
| HDL cholesterol | Breast cancer | mmol/L | 6 |
| Lymphocyte count | Schizophrenia | $10^9$ cells/L | 6 |
| Height | Atrial fibrillation | cm | 7 |
| Vitamin D | Multiple sclerosis | nmol/L | 3 |
| Weight | Myocardial infarction | kg | 6 |
| C-reactive protein | Alzheimer's disease | mg/L | 4 |
| Bone mineral density | Osteoporosis | z-score | 3 |
| Urea | Gout | mmol/L | 6 |
| Basophil counts | Atopic dermatitis | $10^9$ cells/L | 6 |
| Hip circumference | Crohn's disease | cm | 3 |
| Mean corpuscular hemoglobin | Total bilirubin | pg | 3 |
| Mean corpuscular hemoglobin concentration | Iron deficiency anemia | g/dL | 4 |
| Monocyte count | Asthma | $10^9$ cells/L | 6 |
| Neutrophil count | COPD | $10^9$ cells/L | 6 |
| Platelet count | Hypertension | $10^9$ cells/L | 6 |
| RBC count | Total bilirubin | $10^{12}$ cells/L | 6 |
| Triglycerides | Hyperlipidemia | mmol/L | 5 |
| Uric acid | Gout | mg/dL | 6 |
| Waist-hip ratio | Type 2 diabetes | Unitless | 3 |
| Waist circumference | NAFLD/MASLD | cm | 3 |
| WBC count | Coronary artery disease | $10^9$ cells/L | 6 |

Table S1: A summary of the 26 exposure traits we included in our analysis, and the standard unit used for rescaling MR estimates involving the exposure. Each of these exposure traits was linked to a matching outcome trait that we expected it to have a likely non-null effect on, either through literature or via EpiGraphDB[16]. More details on the specific studies for each of these exposure traits is located in Supplemental Data 2.

| Outcome Trait | Number of Studies |
|---|---|
| Atrial Fibrillation | 5 |
| Alzheimer's | 2 |
| Anemia | 4 |
| Asthma | 4 |
| Atopic Dermatitis | 4 |
| Bilirubin | 4 |
| Breast cancer | 4 |
| Coronary Heart Disease | 5 |
| COPD | 6 |
| Crohn's disease | 3 |
| Gout | 6 |
| Hyperlipidemia | 3 |
| Hypertension | 5 |
| Myocardial Infarction | 5 |
| Multiple Sclerosis | 3 |
| NAFLD/MASLD | 2 |
| Osteoporosis | 3 |
| Rheumatoid arthritis | 5 |
| Schizophrenia | 4 |
| Stroke | 5 |
| Type 2 diabetes | 2 |

Table S2: A summary of the 21 outcome traits we included in our analysis, including its standard unit for rescaling and the number of studies for each trait. More details on the specific studies for each of these outcome traits is located in Supplemental Data 3.

| Subpopulation | Scott et al 2016 Equivalent |
|---|---|
| UK Biobank | Great Britain |
| Biobank Japan | Japanese |
| FinnGen | Finnish |
| Taiwan Biobank | Chinese (South) |
| Mixed (majority Hispanic) mega analysis (Wojcik) | No equivalent |
| Pan-UKB South Asians | Persia and Pakistan |
| Pan-UKB Africans | Yoruba (Nigeria) |

Table S3: Closest equivalents of subpopulations in our MR survey to the populations in Scott et al 2016.

| Trait Pair | IV Selection Threshold | Exposure Population | Target Population | Mismatching estimate (p-value) | Reference estimate (p-value) | Concordance z-score |
|---|---|---|---|---|---|---|
| Body Mass Index to Breast Cancer | $5 \times 10^{-8}$ | Biobank Japan | UK Biobank | -0.074 ($p < 0.0001$) | -0.006 ($p = 0.44$) | -3.36 |
| HDL cholesterol to Type 2 diabetes | $1 \times 10^{-5}$ | UK Biobank | Biobank Japan | -0.47 ($p < 0.0001$) | -0.04 ($p = 0.55$) | -4.48 |
| HDL cholesterol to Type 2 diabetes | $5 \times 10^{-8}$ | UK Biobank) | Biobank Japan | -0.41 ($p < 0.0001$) | -0.03 ($p = 0.68$) | -3.40 |
| Height to Atrial Fibrillation | $5 \times 10^{-8}$ | FinnGen | UK Biobank | 0.064 ($p < 0.0001$) | 0.039 ($p < 0.0001$) | 3.35 |
| Weight to Breast Cancer | $5 \times 10^{-8}$ | Taiwan Biobank | UK Biobank | -0.023 ($p = 0.0009$) | -0.001 ($p = 0.60$) | -2.94 |
| Weight to Type 2 diabetes | $5 \times 10^{-8}$ | Taiwan Biobank | Biobank Japan | 0.054 ($p < 0.0001$) | 0.012 ($p = 0.20$) | 2.87 |

Table S4: The concordance z-scores and estimate effect sizes of mismatching MR estimates that are larger in magnitude than their corresponding references in the same direction, and with significant concordance z-scores at an FDR of 5%.

| **MR Method** | **IVW** | **BEE** $(1 \times 10^{-5})$ | **BEE** $(5 \times 10^{-8})$ |
|---|---|---|---|
| Total Number of Mismatching Estimates with Reference Estimates | 1,951 | 2,182 | 1,951 |
| Mismatching Estimates with Non-Significant Concordance z-scores | 1,777 | 1,852 | 1,841 |
| Mismatching Estimates with Significant Concordance z-scores, Same Sign as Reference, Smaller Magnitude | 101 | 227 | 66 |
| Mismatching Estimates with Significant Concordance z-scores, Same Sign as Reference, Larger Magnitude | 7 | 1 | 4 |
| Mismatching Estimates with Significant Concordance z-scores, Opposite Sign as Reference | 66 | 102 | 40 |
| Mismatching Estimates with Significant Concordance z-scores, Opposite Sign as Reference Reference and Mismatch MR p-values $< 0.05$ | 15 | 10 | 13 |
| SIMEX-Estimated Overall Shrinkage Coefficient, 1000 Bootstrap Resamples | 0.453 (SE 0.018) | 0.376 (SE 0.014) | 0.377 (SE 0.017) |

Table S5: A summary of how MR estimates with mismatching exposures compare to their corresponding reference MR estimates for the inverse-variance weighted regression (IVW) and MRBEE methods. MR estimates for IVW were computed at an instrument selection threshold of $5 \times 10^{-8}$, while MR estimates with MRBEE were computed at instrument selection thresholds of $1 \times 10^{-5}$ and $5 \times 10^{-8}$.

| Instrument Selection Threshold | $1 \times 10^{-5}$ | $5 \times 10^{-8}$ |
|---|---|---|
| Total Number of Mismatching Estimates with Reference Estimates | 2,182 | 1,951 |
| Mismatching Estimates with Non-Significant Concordance z-scores | 1,772 | 1,792 |
| Mismatching Estimates with Significant Concordance z-scores, Same Sign as Reference, Smaller Magnitude | 287 | 102 |
| Mismatching Estimates with Significant Concordance z-scores, Same Sign as Reference, Larger Magnitude | 3 | 5 |
| Mismatching Estimates with Significant Concordance z-scores, Opposite Sign as Reference | 120 | 52 |
| Mismatching Estimates with Significant Concordance z-scores, Opposite Sign as Reference, Reference and Mismatch MR p-values $< 0.05$ | 6 | 8 |

Table S6: A summary of how MR estimates with mismatching exposures compared with their corresponding reference MR estimates at two different instrument selection thresholds. All estimates were computed using GRAPPLE, but without the Steiger filtering of the main analysis.

| Trait Pair | Significance of Meta-analysis and BBJ Exposures | Populations Used in Replication | Replication Status |
|---|---|---|---|
| Weight to Coronary Heart Disease | Meta-analysis Only | FinnGen | Replicated |
| Hemoglobin A1c to Coronary Heart Disease | Meta-analysis Only | | Not Tested |
| Monocyte count to Bilirubin | Meta-analysis Only | AFR (Pan-UKB), SAS (Pan-UKB) | Not Replicated |
| RBC count to Bilirubin | Meta-analysis Only | AFR (Pan-UKB), SAS (Pan-UKB) | Replicated |
| Hemoglobin A1c to Bilirubin | Meta-analysis Only | | Not Tested |
| Height to COPD | Meta-analysis Only | FinnGen | Not Replicated |
| RBC count to Myocardial Infarction | Meta-analysis Only | | Not Tested |
| Height to Myocardial Infarction | Meta-analysis Only | FinnGen | Replicated |
| Weight to Myocardial Infarction | Meta-analysis Only | FinnGen | Replicated |
| Hemoglobin A1c to Myocardial Infarction | Meta-analysis Only | | Not Tested |
| Weight to Stroke | Meta-analysis Only | FinnGen | Replicated |
| C-reactive protein to Type 2 diabetes | Meta-analysis Only | Majority Hispanic mega-analysis | Replicated |
| HDL cholesterol to Type 2 diabetes | Meta-analysis Only | Majority Hispanic mega-analysis | Replicated |
| Triglycerides to Type 2 diabetes | Meta-analysis Only | Majority Hispanic mega-analysis | Replicated |
| HDL cholesterol to Bilirubin | BBJ Only | | Not Tested |
| Height to Asthma | BBJ Only | FinnGen | Replicated |
| Triglycerides to COPD | BBJ Only | | Not Tested |
| Weight to Atrial Fibrillation | BBJ Only | FinnGen | Replicated |
| Height to Type 2 diabetes | BBJ Only | Majority Hispanic mega-analysis, FinnGen | Replicated |

Table S7: Replication analysis results in non-UK and non-Japanese populations for 19 causal effect estimates that differed in significance ($p < 0.05/195$) when the exposure was measured either only in Biobank Japan or in a meta analysis of UK Biobank and Biobank Japan using instrument selection threshold $10^{-5}$. An effect was considered replicated if it had a non-UK, non-Japanese MR estimate in the same direction as the significant estimate, with a p-value below a Bonferroni-corrected threshold (0.05/13).

| Trait Pair | Significance of Meta analysis and BBJ Exposures | Populations Used in Replication | Replication Status |
|---|---|---|---|
| Height to Coronary Heart Disease | Meta analysis Only | FinnGen | Replicated |
| Hemoglobin A1c to Coronary Heart Disease | Meta analysis Only | | Not Tested |
| Monocyte count to Bilirubin | Meta analysis Only | AFR (Pan UKB), SAS (Pan UKB) | Not Replicated |
| RBC count to Bilirubin | Meta analysis Only | AFR (Pan UKB), SAS (Pan UKB) | Not Replicated |
| Hemoglobin A1c to Bilirubin | Meta analysis Only | | Not Tested |
| Height to Asthma | Meta analysis Only | FinnGen | Replicated |
| Height to COPD | Meta analysis Only | FinnGen | Not Replicated |
| Height to Myocardial Infarction | Meta analysis Only | FinnGen | Replicated |
| Weight to Myocardial Infarction | Meta analysis Only | FinnGen | Replicated |
| Hemoglobin A1c to Myocardial Infarction | Meta analysis Only | | Not Tested |
| Platelet count to Type 2 diabetes | Meta analysis Only | Majority Hispanic mega analysis | Not Replicated |
| Weight to Type 2 diabetes | Meta analysis Only | FinnGen | Replicated |
| HDL cholesterol to Type 2 diabetes | Meta analysis Only | Majority Hispanic mega analysis | Not Replicated |
| Hemoglobin A1c to Type 2 diabetes | Meta analysis Only | Majority Hispanic mega analysis | Not Replicated |
| Triglycerides to COPD | BBJ Only | | Not Tested |
| Weight to Atrial Fibrillation | BBJ Only | FinnGen | Replicated |

Table S8: Replication analysis results in non-UK and non-Japanese populations for 19 causal effect estimates that differed in significance ($p < 0.05/195$) when the exposure was measured either only in Biobank Japan or in a meta analysis of UK Biobank and Biobank Japan using instrument selection threshold $5 \times 10^{-8}$. An effect was considered replicated if it had a non-UK, non-Japanese MR estimate in the same direction as the significant estimate, with a p-value below a Bonferroni-corrected threshold (0.05/12).

#### 6 Supplemental Figures

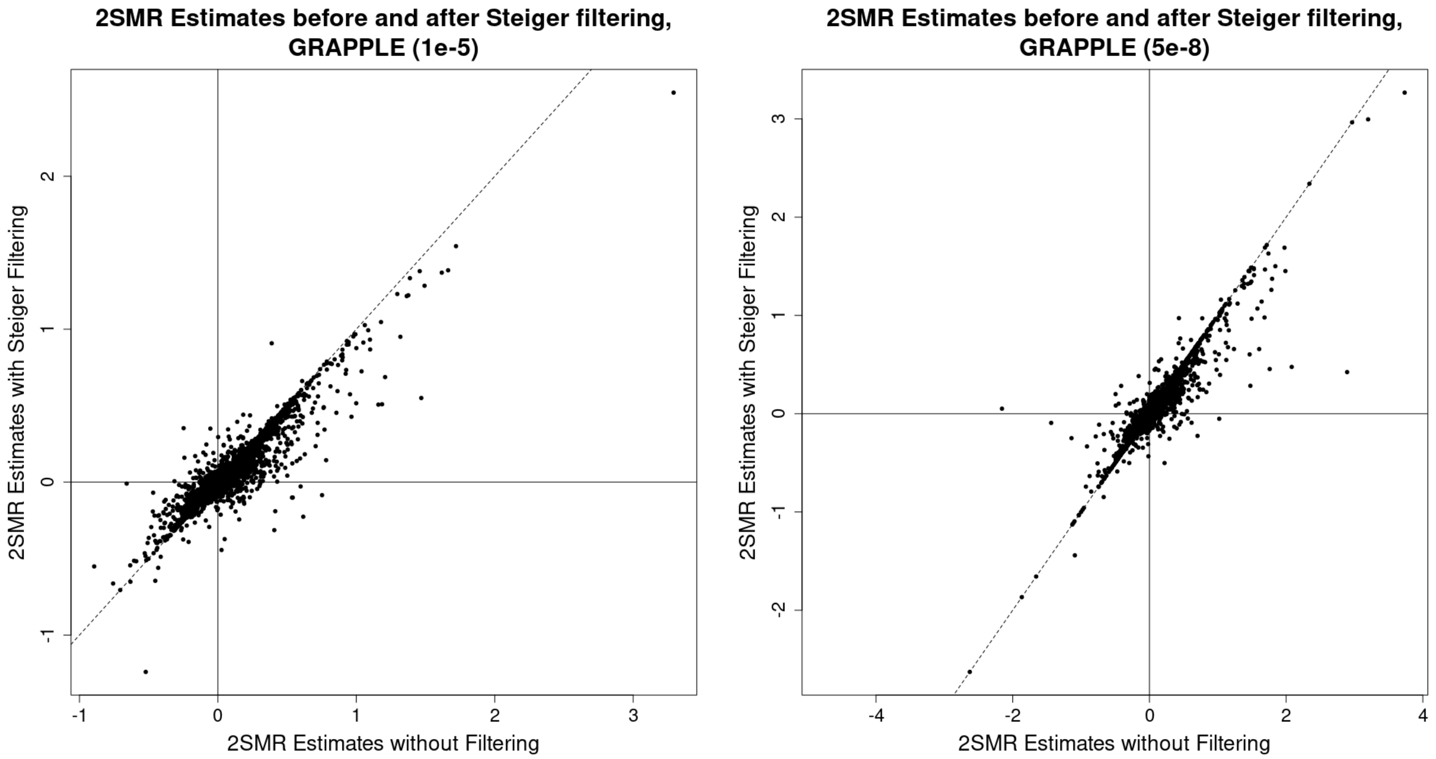


Figure S1: A comparison of MR estimates estimated using GRAPPLE before and after Steiger filtering at two instrument selection thresholds. The dashed line represents the $y = x$ line. All estimates were not yet rescaled to the respective standard units of their exposure.

A) GRAPPLE, $1 \times 10^{-5}$

| Target → Exposure ↓ | UK Biobank | FinnGen | Biobank Japan | Taiwan Biobank | Hispanic mega-analysis | SAS (UK) | AFR (UK) |
|---|---|---|---|---|---|---|---|
| **UK Biobank** | | 0.67 (0.02) [40] | 0.44 (0.03) [260] | 0.15 (0.09) [9] | 0.68 (0.17) [12] | 0.11 (0.05) [40] | 0.02 (0.13) [8] |
| **FinnGen** | 0.53 (0.02) [36] | | 0.50 (0.05) [26] | | | | |
| **Biobank Japan** | 0.25 (0.02) [360] | 0.32 (0.02) [40] | | | 0.23 (0.14) [12] | 0.14 (0.08) [40] | -0.32 (0.25) [8] |
| **Taiwan Biobank** | 0.40 (0.02) [162] | 0.34 (0.04) [20] | 0.77 (0.04) [91] | | | | |
| **Hispanic mega-analysis** | 0.21 (0.02) [108] | 0.15 (0.02) [40] | 0.36 (0.04) [78] | | | 0.30 (0.24) [8] | |
| **SAS (UK)** | 0.06 (0.02) [180] | | 0.06 (0.03) [130] | | | | -0.34 (0.19) [8] |
| **AFR (UK)** | -0.01 (0.01) [144] | | -0.01 (0.02) [104] | | | 0.07 (0.03) [32] | |

B) GRAPPLE, $5 \times 10^{-8}$

| Target → Exposure ↓ | UK Biobank | FinnGen | Biobank Japan | Taiwan Biobank | Hispanic mega-analysis | SAS (UK) | AFR (UK) |
|---|---|---|---|---|---|---|---|
| **UK Biobank** | | 0.70 (0.03) [40] | 0.26 (0.03) [260] | 0.14 (0.09) [9] | 0.40 (0.11) [12] | 0.04 (0.03) [36] | |
| **FinnGen** | 0.65 (0.03) [36] | | 0.53 (0.05) [26] | | | | |
| **Biobank Japan** | 0.31 (0.02) [360] | 0.39 (0.03) [40] | | | 0.29 (0.14) [12] | 0.14 (0.05) [36] | |
| **Taiwan Biobank** | 0.56 (0.03) [162] | 0.38 (0.06) [20] | 0.67 (0.05) [91] | | | | |
| **Hispanic mega-analysis** | 0.52 (0.03) [108] | 0.47 (0.04) [40] | 0.57 (0.05) [78] | | | 0.12 (0.14) [8] | |
| **SAS (UK)** | 0.02 (0.10) [124] | | 0.04 (0.05) [87] | | | | |
| **AFR (UK)** | 0.01 (0.02) [108] | | -0.03 (0.02) [78] | | | 0.03 (0.02) [24] | |

Figure S2: Population pair-specific SIMEX-estimated shrinkage coefficients. Bootstrap standard error is given in round parentheses. The number of estimates contributing to each coefficient is given in square parentheses. No estimate was computed when fewer than eight estimates for a population pair were available.

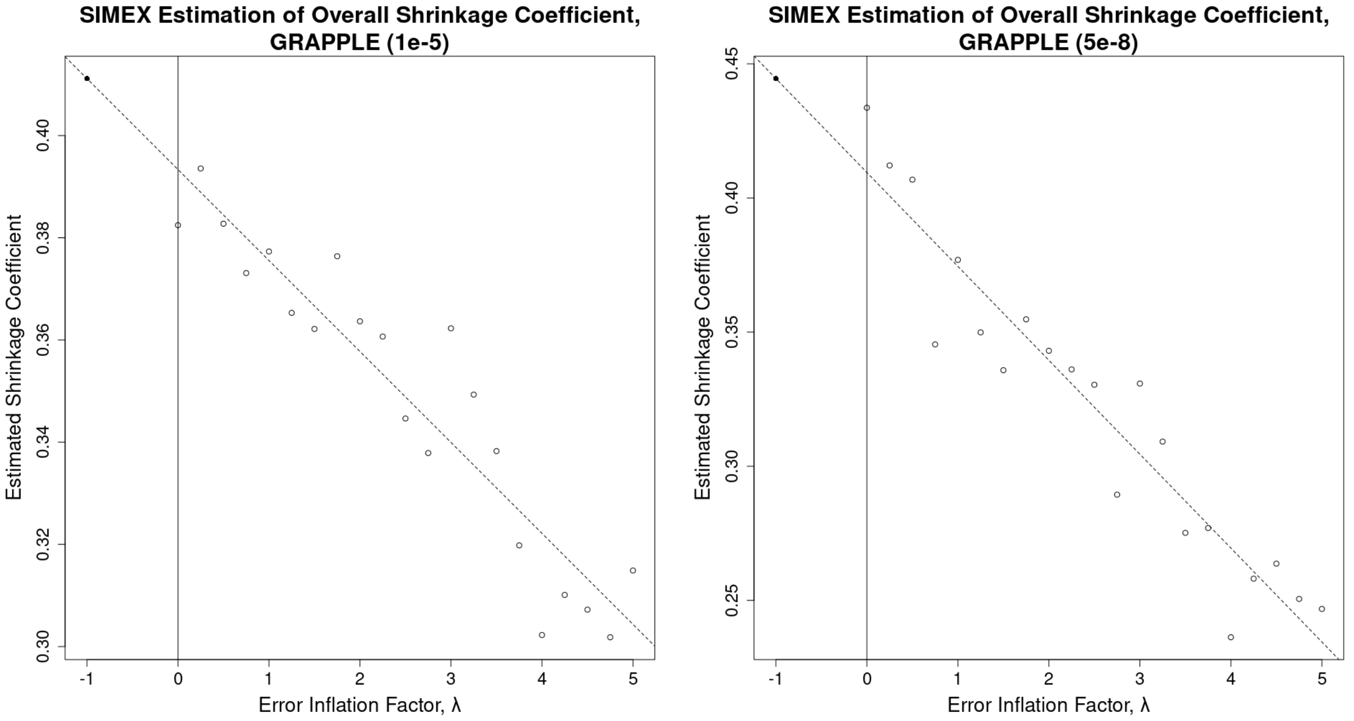


Figure S3: Estimated overall shrinkage coefficients of MR vs additional measurement error (SIMEX $\lambda$) for two instrument selection thresholds. Estimated shrinkage coefficients with added noise are shown as open points. The SIMEX-estimated "noise-free" coefficient at $\lambda = -1$ is extrapolated using the noisier estimates, and is shown as a solid point. The slope and intercept of the linear regression used for extrapolation is shown as a dashed line.

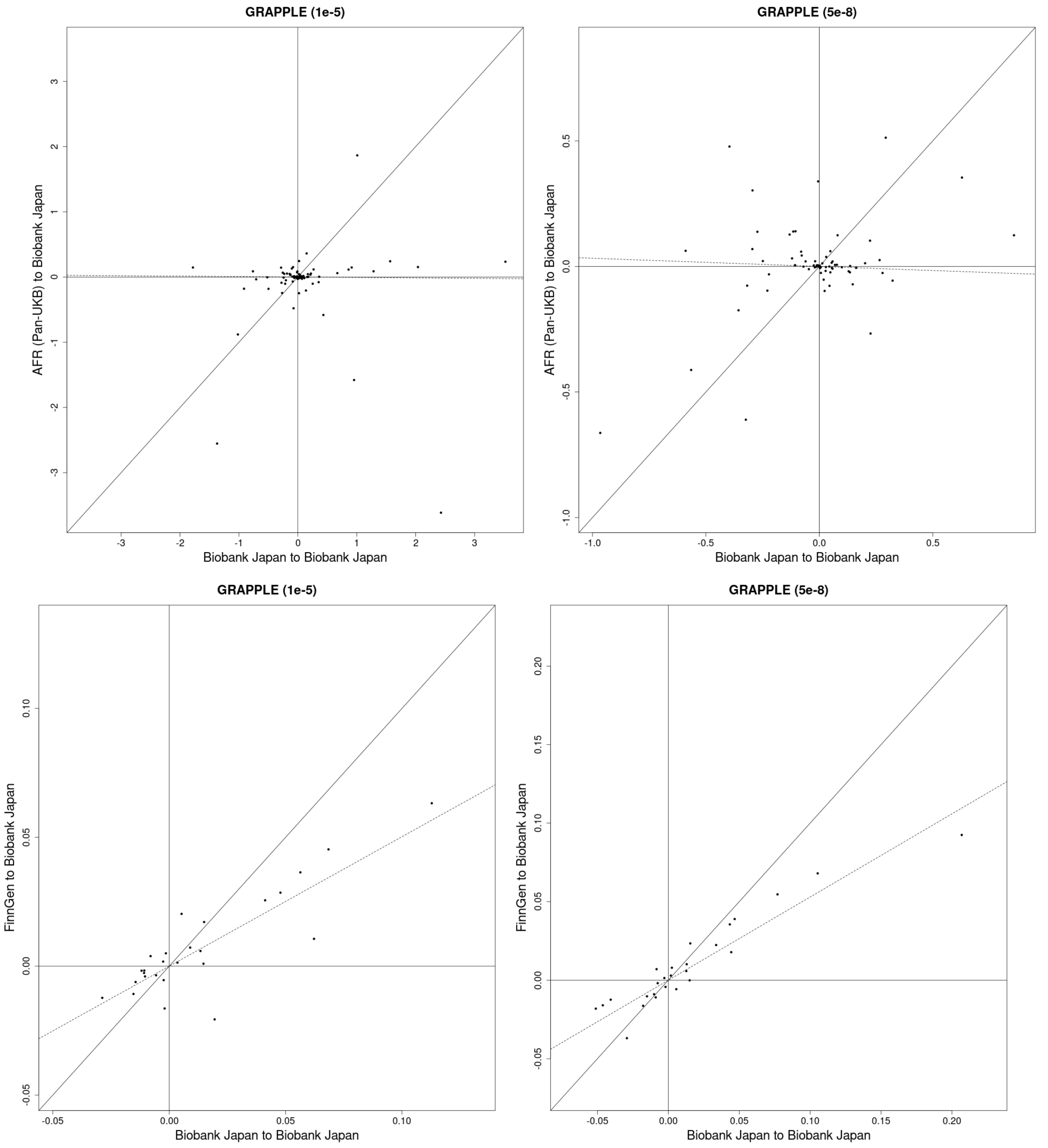
GRAPPLE (1e-5)
AFR (Pan-UKB) to Biobank Japan
Biobank Japan to Biobank Japan
GRAPPLE (5e-8)
AFR (Pan-UKB) to Biobank Japan
Biobank Japan to Biobank Japan
GRAPPLE (1e-5)
FinnGen to Biobank Japan
Biobank Japan to Biobank Japan
GRAPPLE (5e-8)
FinnGen to Biobank Japan
Biobank Japan to Biobank Japan

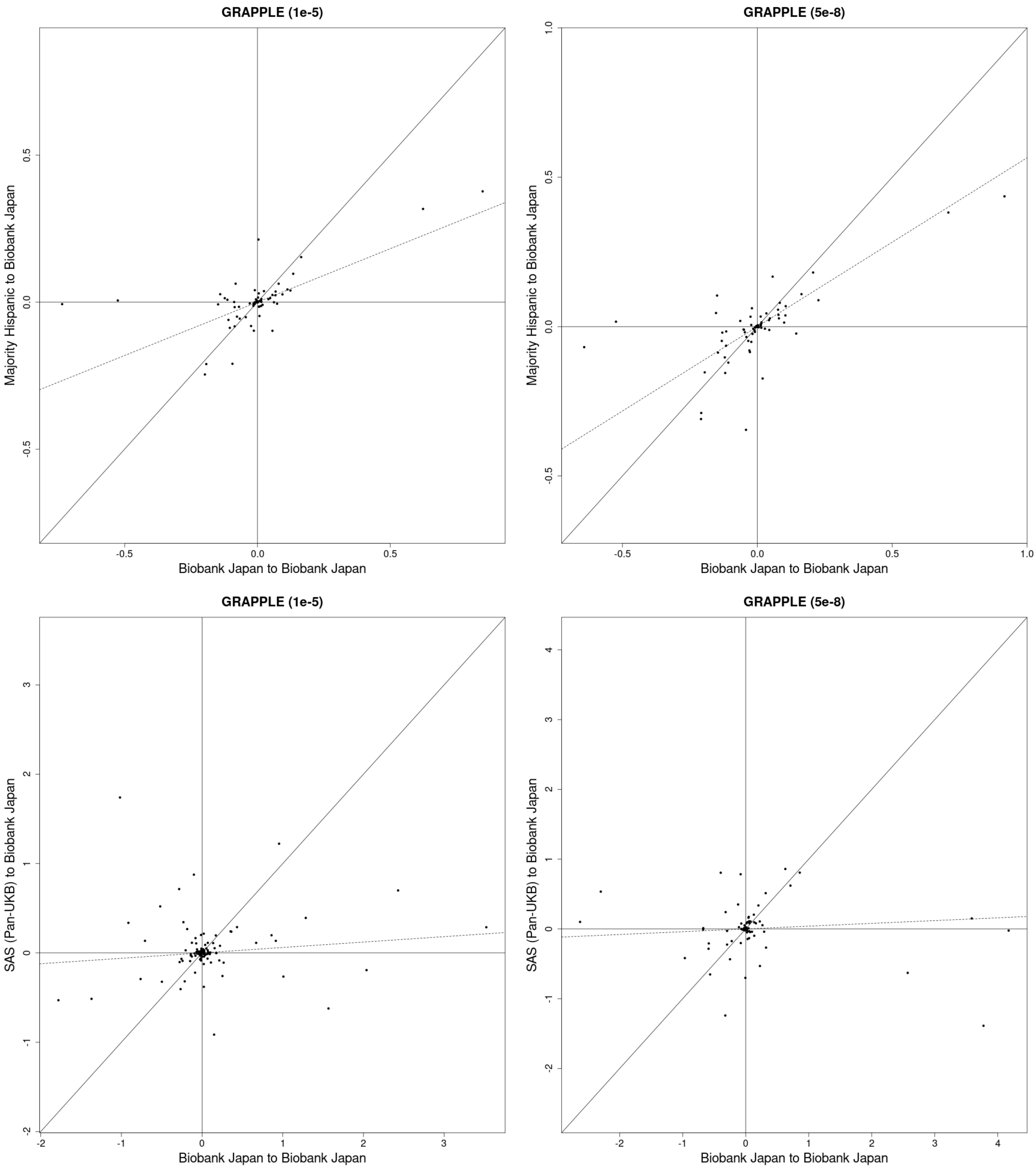
GRAPPLE (1e-5)
Majority Hispanic to Biobank Japan
Biobank Japan to Biobank Japan
-0.5
0.0
0.5
GRAPPLE (5e-8)
Majority Hispanic to Biobank Japan
Biobank Japan to Biobank Japan
-0.5
0.0
0.5
1.0
GRAPPLE (1e-5)
SAS (Pan-UKB) to Biobank Japan
Biobank Japan to Biobank Japan
-2
-1
0
1
2
3
GRAPPLE (5e-8)
SAS (Pan-UKB) to Biobank Japan
Biobank Japan to Biobank Japan
-2
-1
0
1
2
3
4

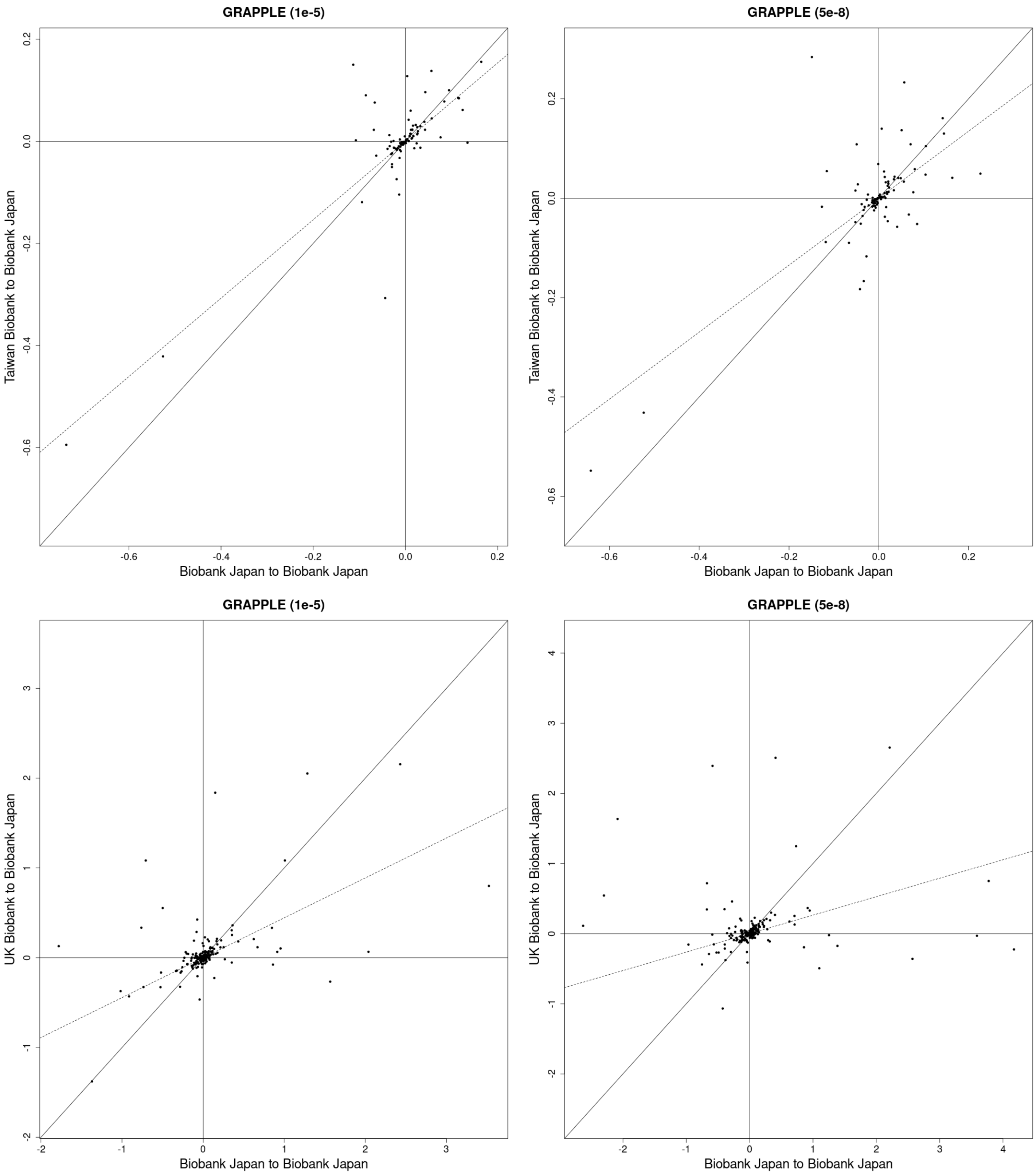
GRAPPLE (1e-5)
Taiwan Biobank to Biobank Japan
Biobank Japan to Biobank Japan
GRAPPLE (5e-8)
Taiwan Biobank to Biobank Japan
Biobank Japan to Biobank Japan
GRAPPLE (1e-5)
UK Biobank to Biobank Japan
Biobank Japan to Biobank Japan
GRAPPLE (5e-8)
UK Biobank to Biobank Japan
Biobank Japan to Biobank Japan

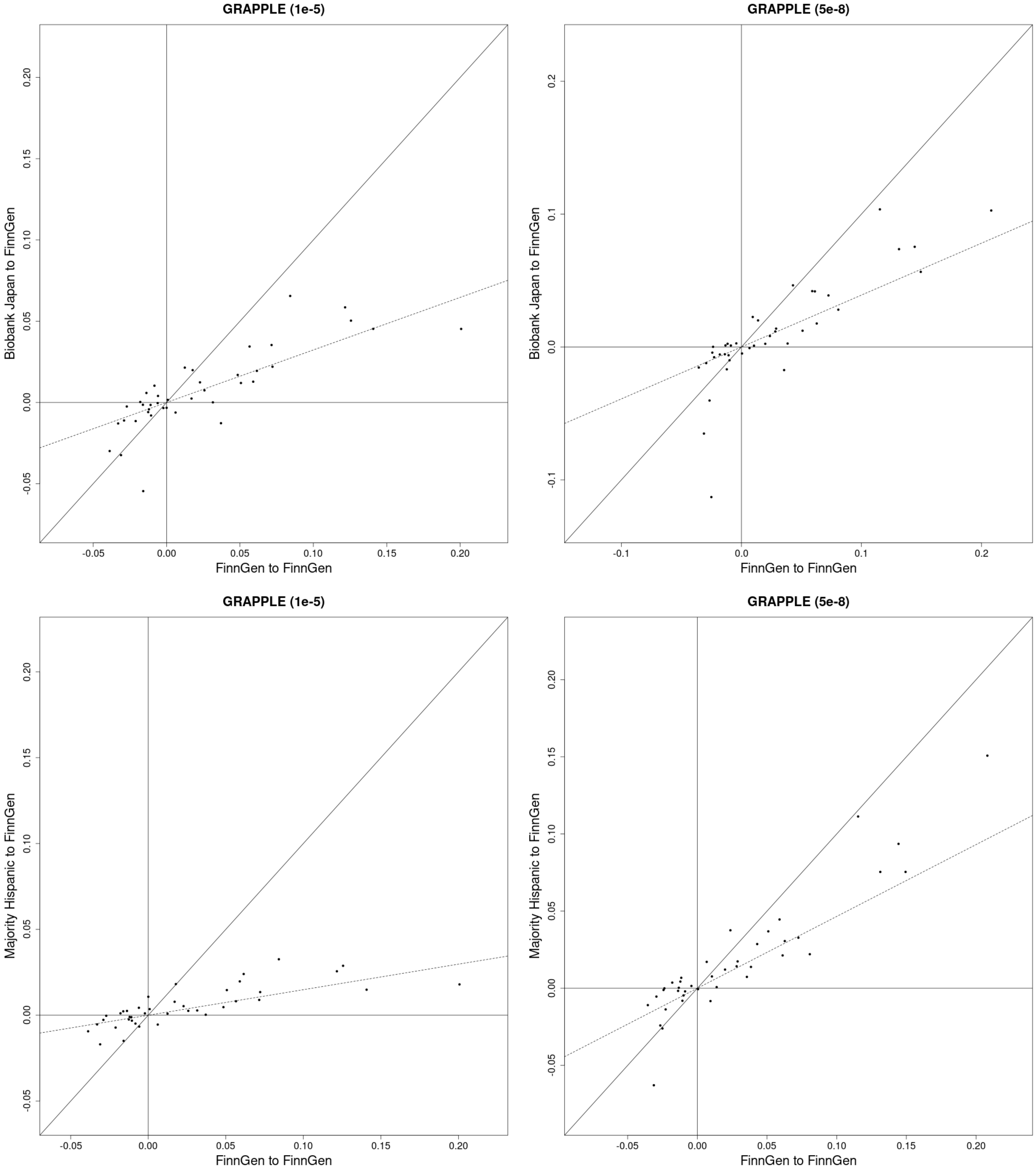
GRAPPLE (1e-5)
Biobank Japan to FinnGen
FinnGen to FinnGen
GRAPPLE (5e-8)
Biobank Japan to FinnGen
FinnGen to FinnGen
GRAPPLE (1e-5)
Majority Hispanic to FinnGen
FinnGen to FinnGen
GRAPPLE (5e-8)
Majority Hispanic to FinnGen
FinnGen to FinnGen

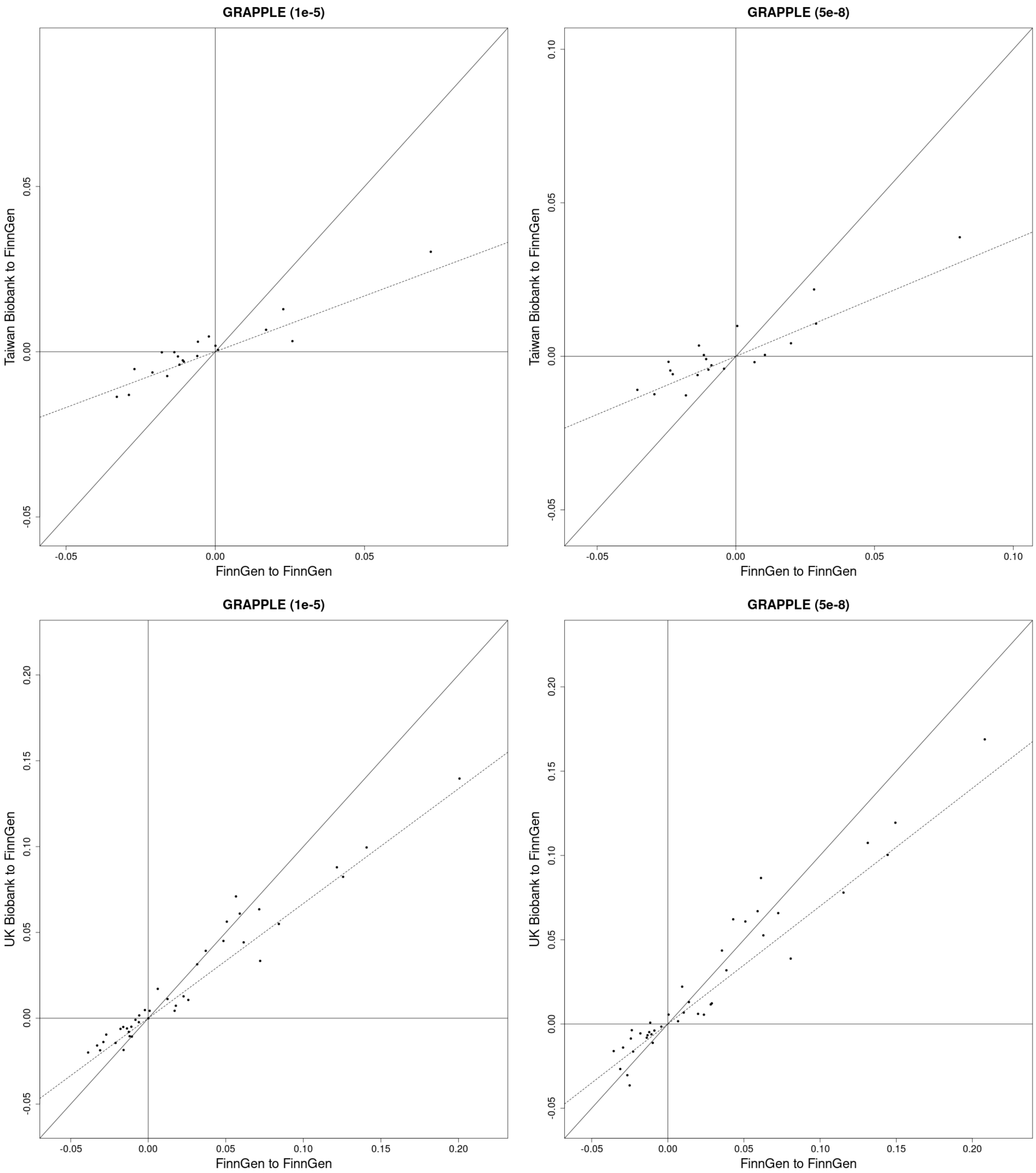
GRAPPLE (1e-5)
Taiwan Biobank to FinnGen
FinnGen to FinnGen
-0.05
0.00
0.05
GRAPPLE (5e-8)
Taiwan Biobank to FinnGen
FinnGen to FinnGen
-0.05
0.00
0.05
0.10
GRAPPLE (1e-5)
UK Biobank to FinnGen
FinnGen to FinnGen
-0.05
0.00
0.05
0.10
0.15
0.20
GRAPPLE (5e-8)
UK Biobank to FinnGen
FinnGen to FinnGen
-0.05
0.00
0.05
0.10
0.15
0.20

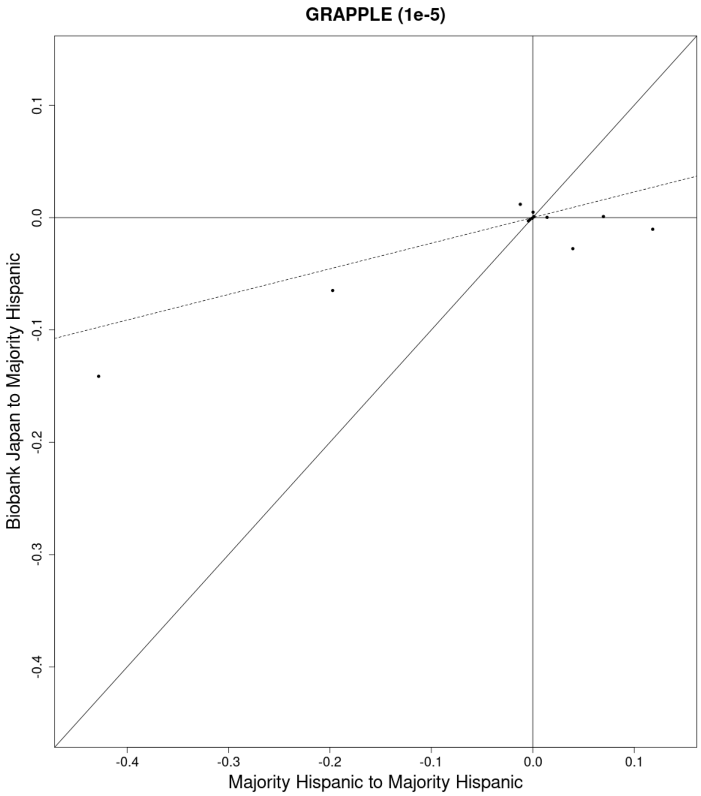
GRAPPLE (1e-5)
Biobank Japan to Majority Hispanic
Majority Hispanic to Majority Hispanic
0.1
0.0
-0.1
-0.2
-0.3
-0.4
-0.4
-0.3
-0.2
-0.1
0.0
0.1

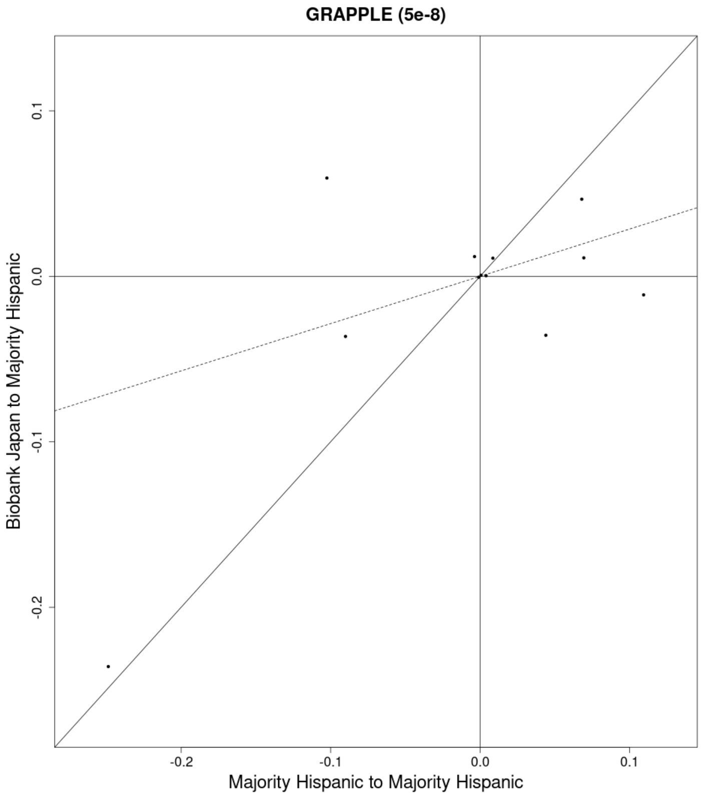
GRAPPLE (5e-8)
Biobank Japan to Majority Hispanic
Majority Hispanic to Majority Hispanic
0.1
0.0
-0.1
-0.2
-0.2
-0.1
0.0
0.1

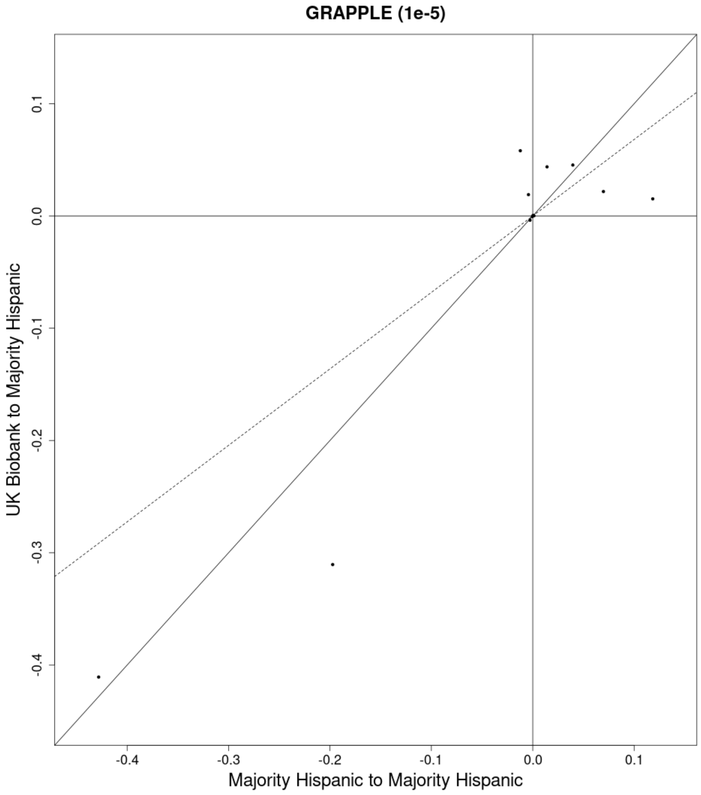
GRAPPLE (1e-5)
UK Biobank to Majority Hispanic
Majority Hispanic to Majority Hispanic
0.1
0.0
-0.1
-0.2
-0.3
-0.4
-0.4
-0.3
-0.2
-0.1
0.0
0.1

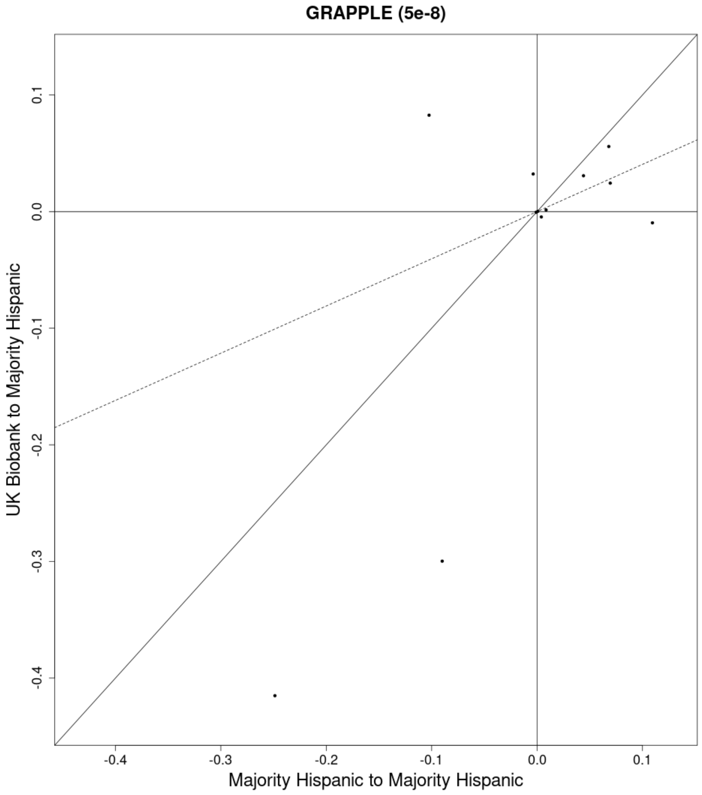
GRAPPLE (5e-8)
UK Biobank to Majority Hispanic
Majority Hispanic to Majority Hispanic
0.1
0.0
-0.1
-0.2
-0.3
-0.4
-0.4
-0.3
-0.2
-0.1
0.0
0.1

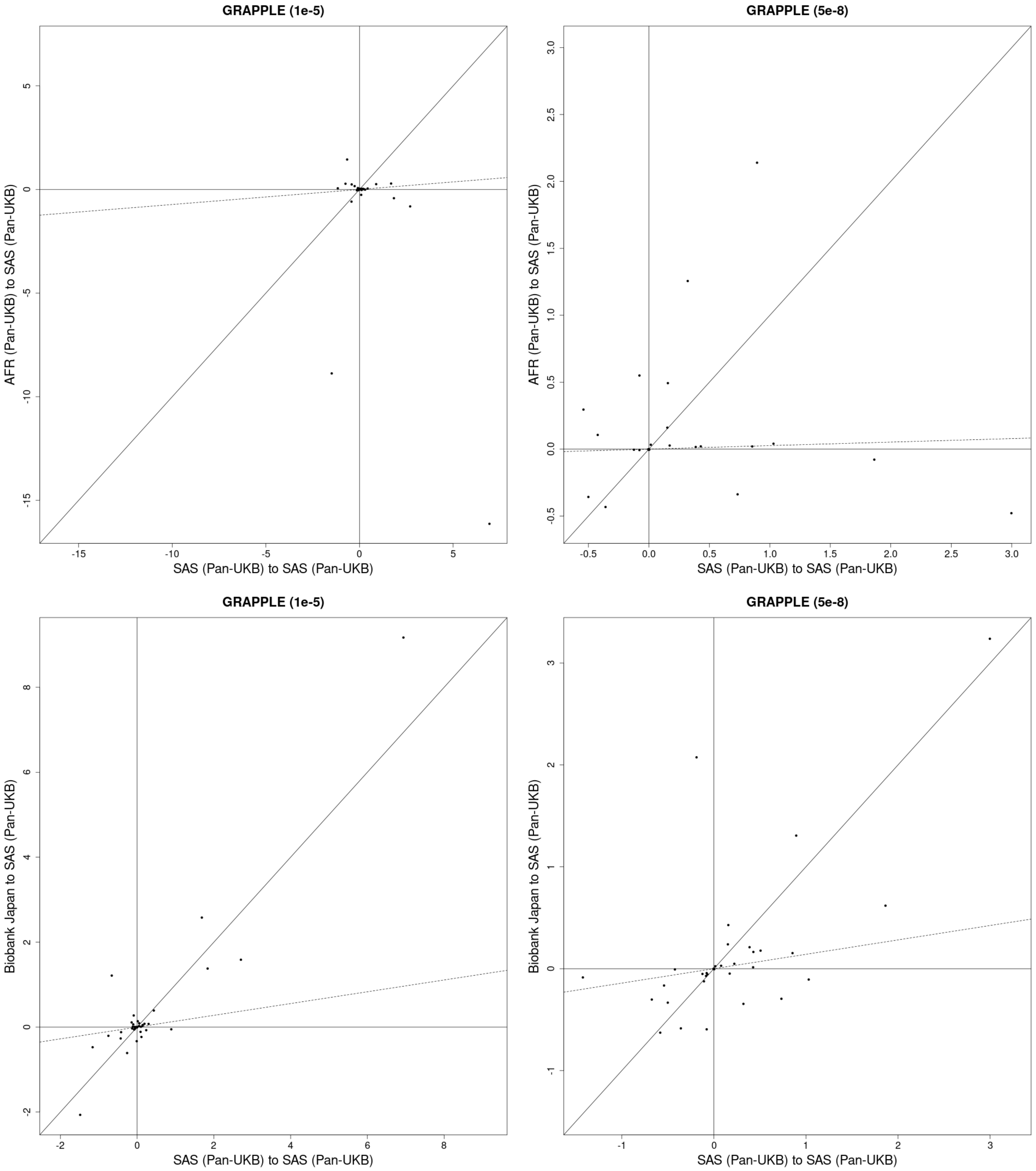
GRAPPLE (1e-5)
AFR (Pan-UKB) to SAS (Pan-UKB)
SAS (Pan-UKB) to SAS (Pan-UKB)
GRAPPLE (5e-8)
AFR (Pan-UKB) to SAS (Pan-UKB)
SAS (Pan-UKB) to SAS (Pan-UKB)
GRAPPLE (1e-5)
Biobank Japan to SAS (Pan-UKB)
SAS (Pan-UKB) to SAS (Pan-UKB)
GRAPPLE (5e-8)
Biobank Japan to SAS (Pan-UKB)
SAS (Pan-UKB) to SAS (Pan-UKB)

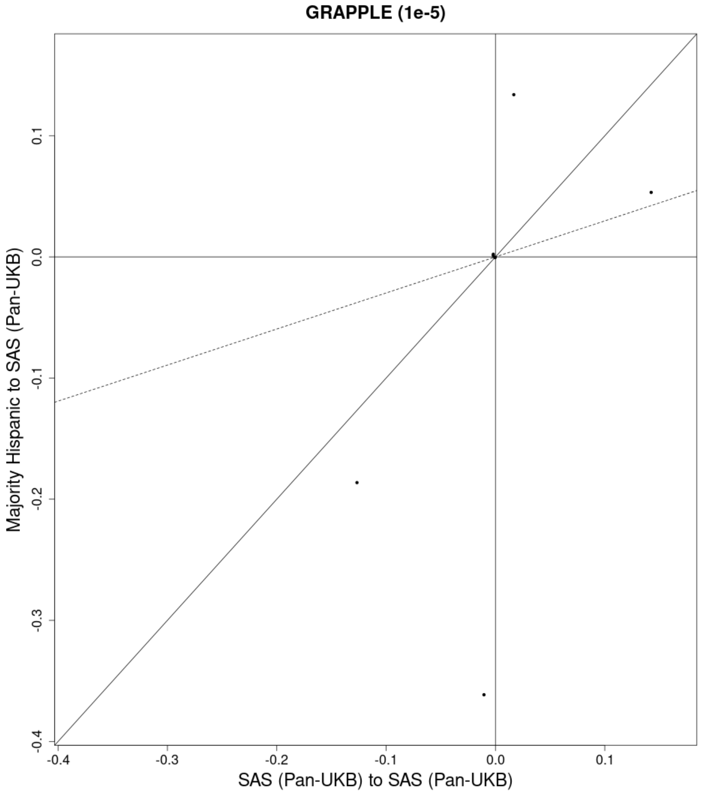
GRAPPLE (1e-5)
Majority Hispanic to SAS (Pan-UKB)
SAS (Pan-UKB) to SAS (Pan-UKB)
-0.4
-0.3
-0.2
-0.1
0.0
0.1

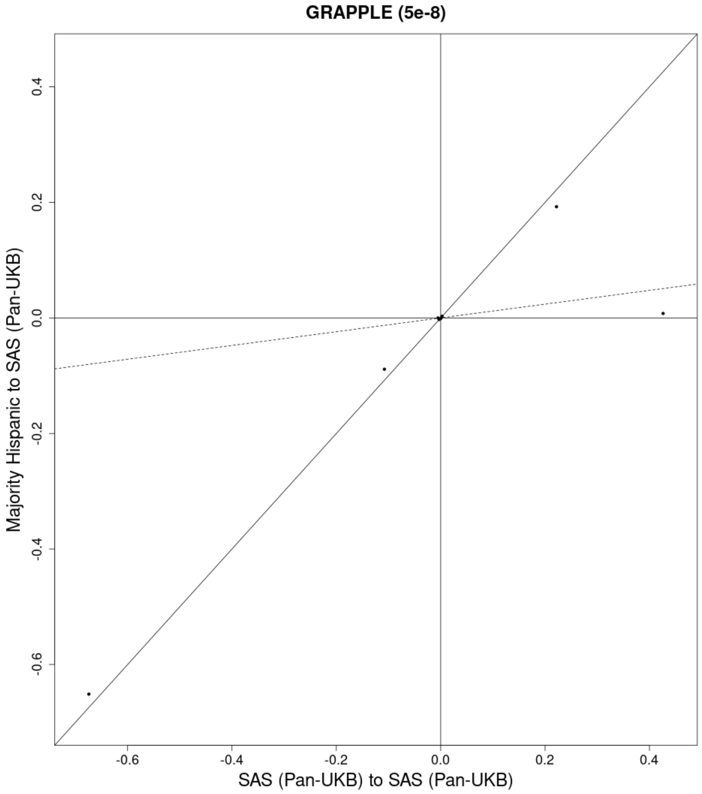
GRAPPLE (5e-8)
Majority Hispanic to SAS (Pan-UKB)
SAS (Pan-UKB) to SAS (Pan-UKB)
-0.6
-0.4
-0.2
0.0
0.2
0.4

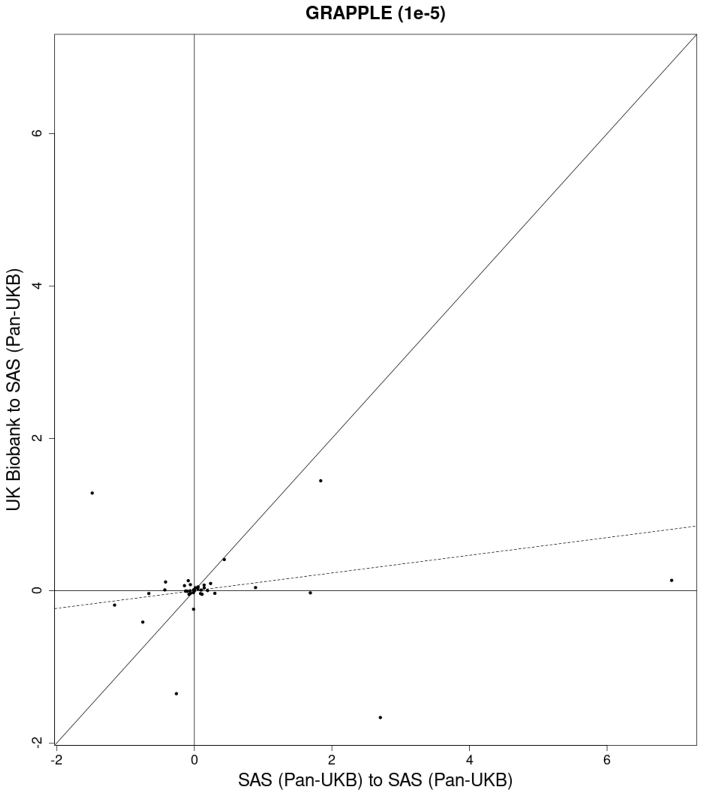
GRAPPLE (1e-5)
UK Biobank to SAS (Pan-UKB)
SAS (Pan-UKB) to SAS (Pan-UKB)
-2
0
2
4
6

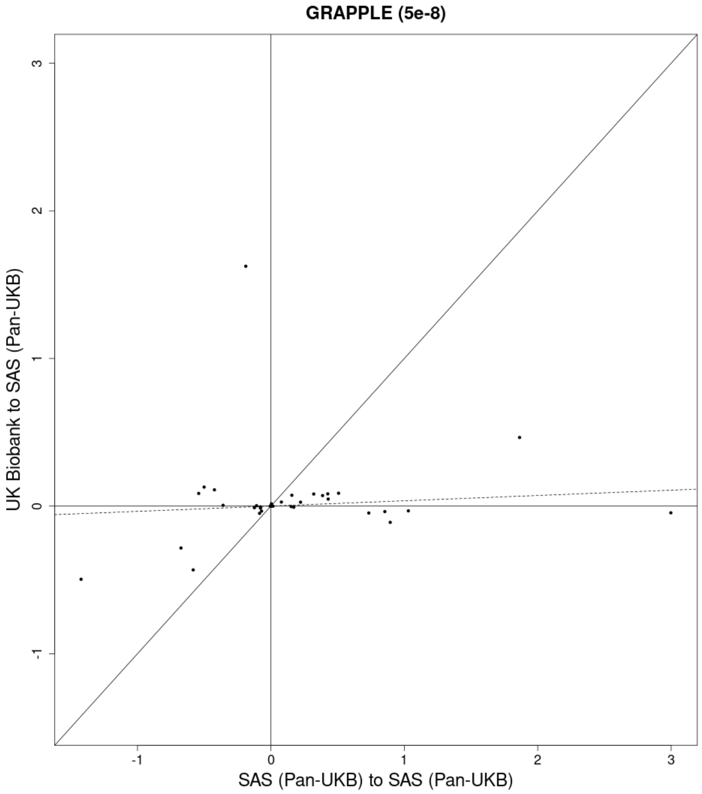
GRAPPLE (5e-8)
UK Biobank to SAS (Pan-UKB)
SAS (Pan-UKB) to SAS (Pan-UKB)
-1
0
1
2
3

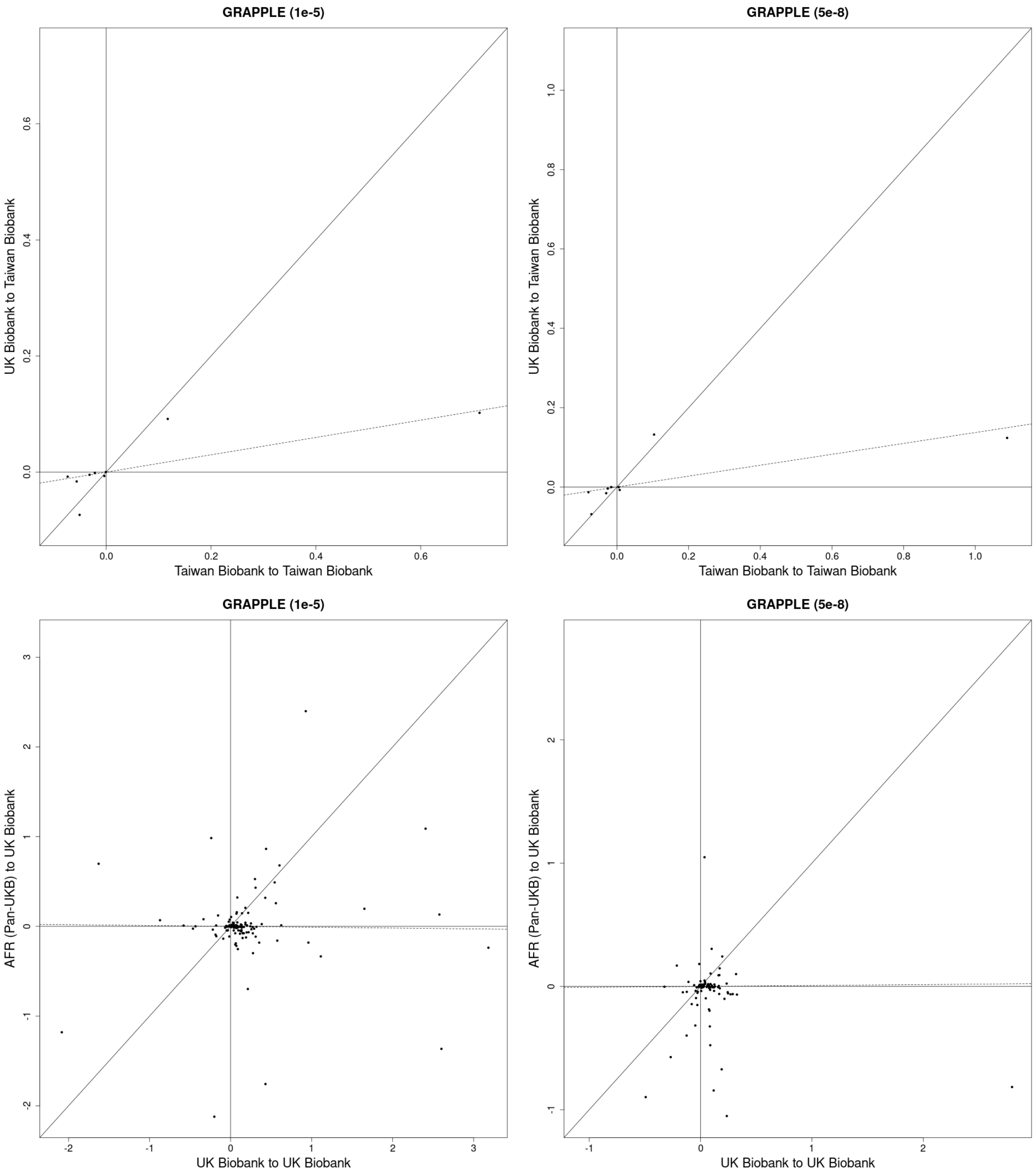
GRAPPLE (1e-5)
UK Biobank to Taiwan Biobank
Taiwan Biobank to Taiwan Biobank
GRAPPLE (5e-8)
UK Biobank to Taiwan Biobank
Taiwan Biobank to Taiwan Biobank
GRAPPLE (1e-5)
AFR (Pan-UKB) to UK Biobank
UK Biobank to UK Biobank
GRAPPLE (5e-8)
AFR (Pan-UKB) to UK Biobank
UK Biobank to UK Biobank

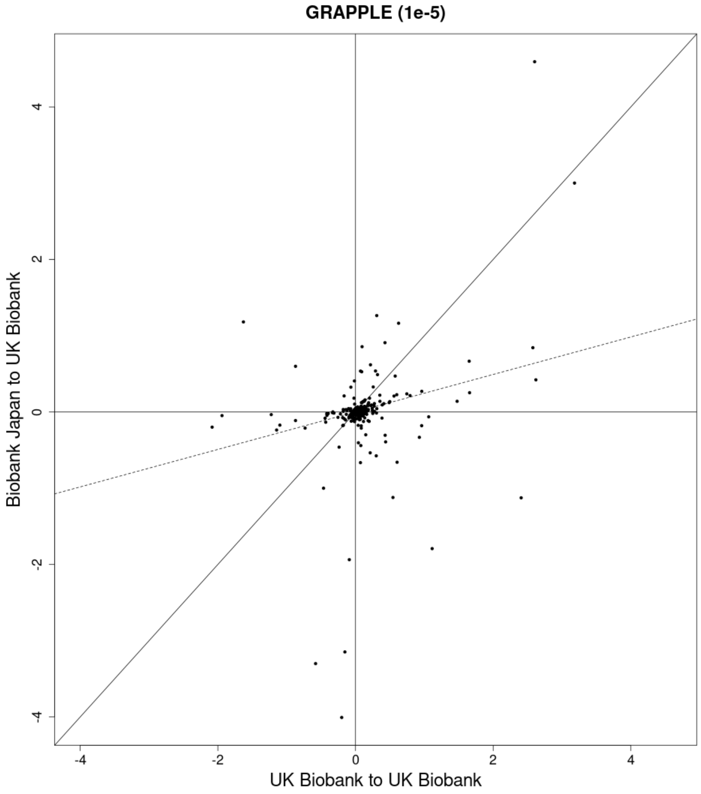
GRAPPLE (1e-5)
Biobank Japan to UK Biobank
UK Biobank to UK Biobank

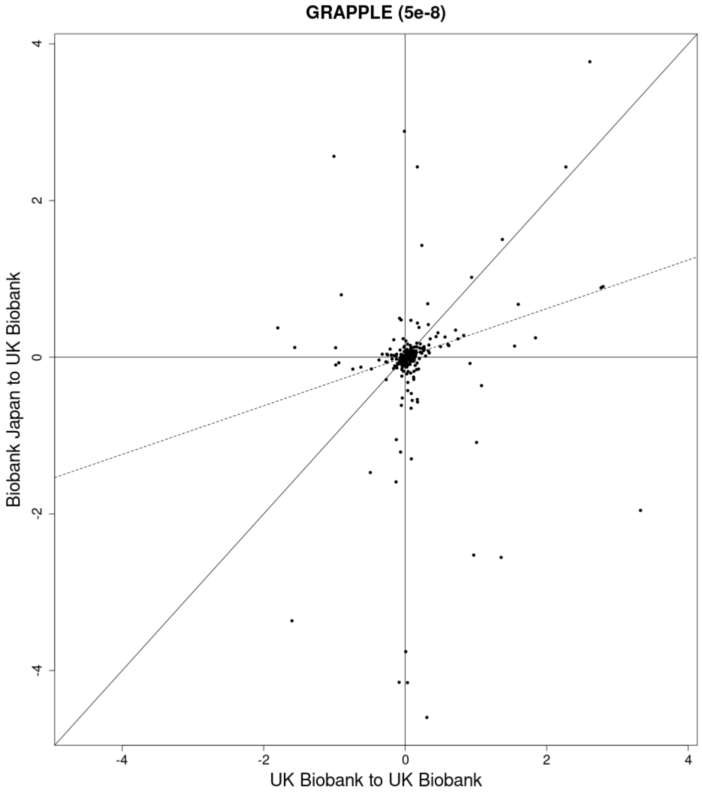
GRAPPLE (5e-8)
Biobank Japan to UK Biobank
UK Biobank to UK Biobank

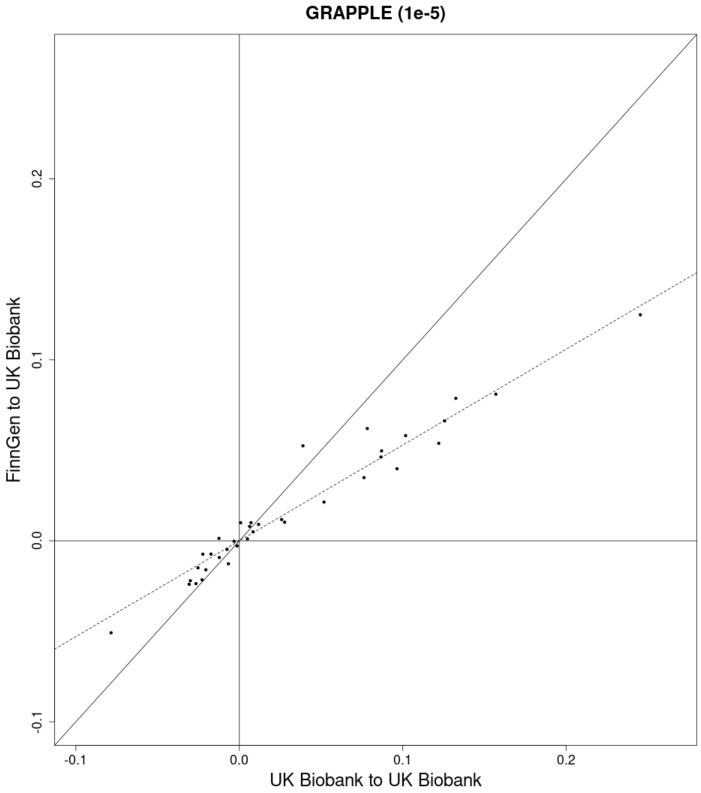
GRAPPLE (1e-5)
FinnGen to UK Biobank
UK Biobank to UK Biobank

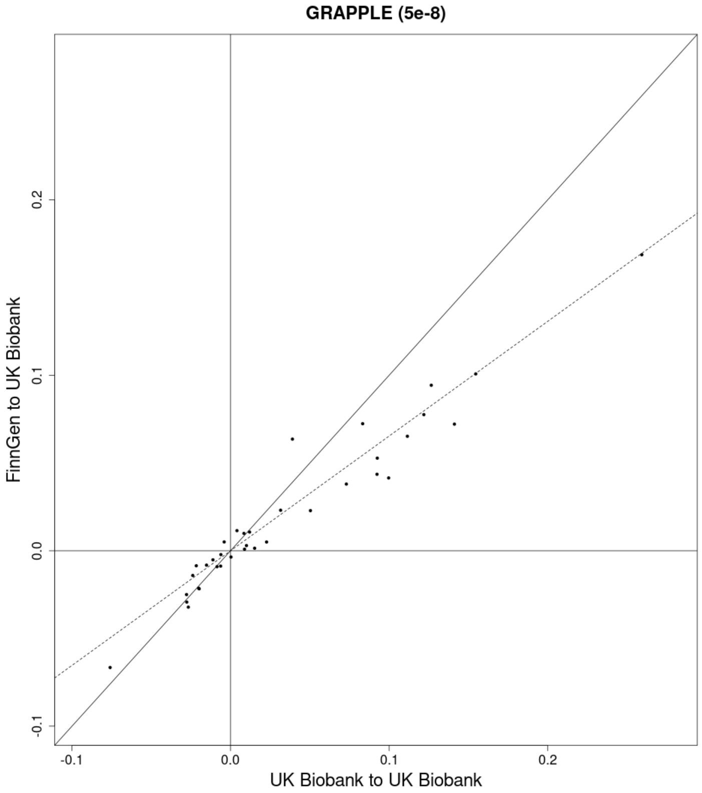
GRAPPLE (5e-8)
FinnGen to UK Biobank
UK Biobank to UK Biobank

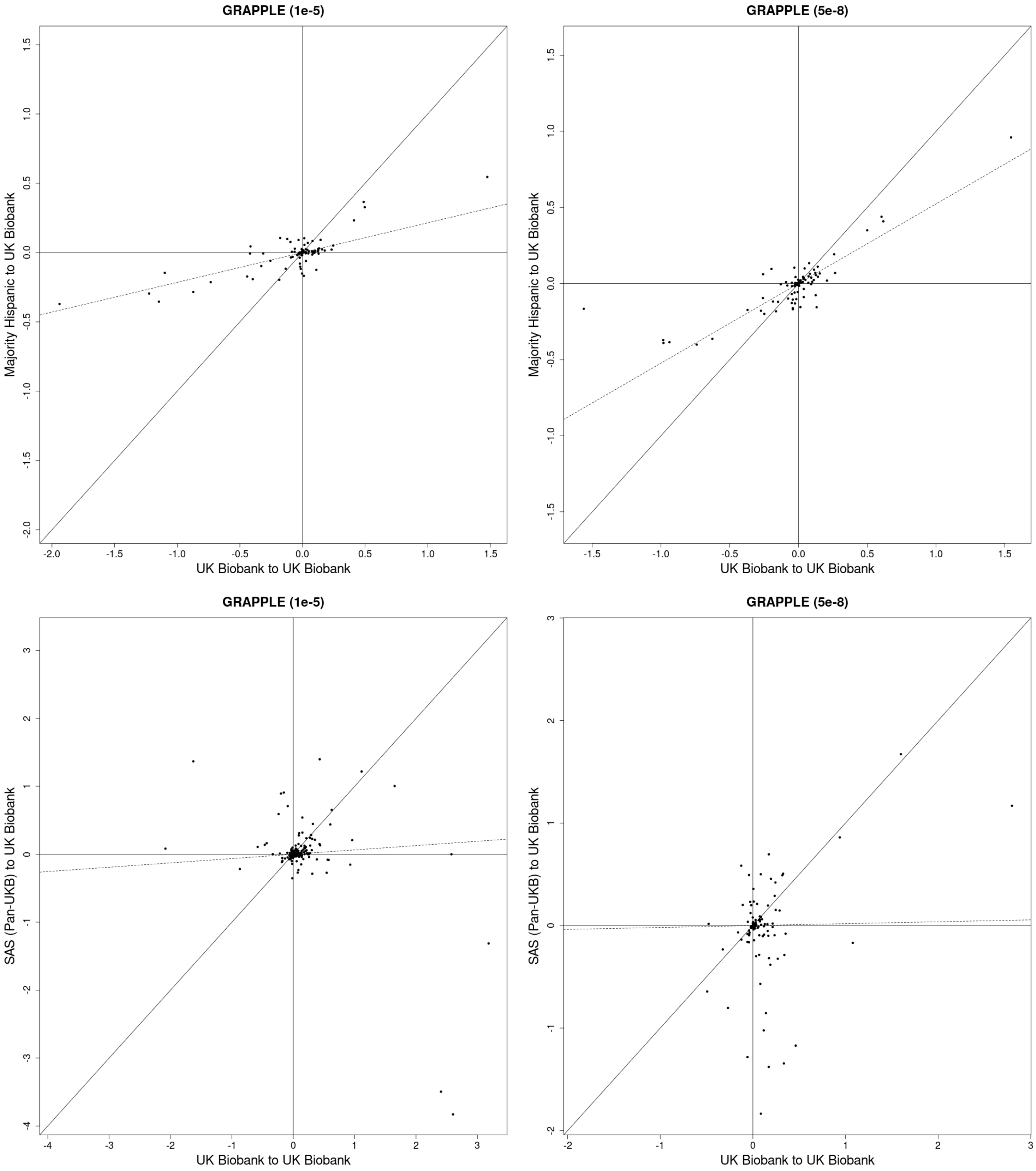
GRAPPLE (1e-5)
Majority Hispanic to UK Biobank
UK Biobank to UK Biobank
GRAPPLE (5e-8)
Majority Hispanic to UK Biobank
UK Biobank to UK Biobank
GRAPPLE (1e-5)
SAS (Pan-UKB) to UK Biobank
UK Biobank to UK Biobank
GRAPPLE (5e-8)
SAS (Pan-UKB) to UK Biobank
UK Biobank to UK Biobank

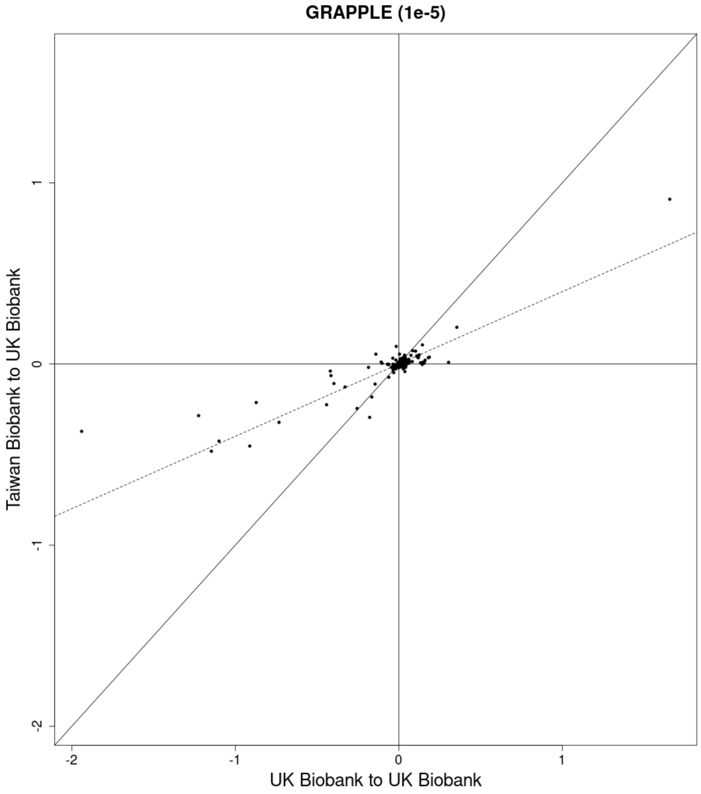
GRAPPLE (1e-5)
Taiwan Biobank to UK Biobank
UK Biobank to UK Biobank
-2
-1
0
1

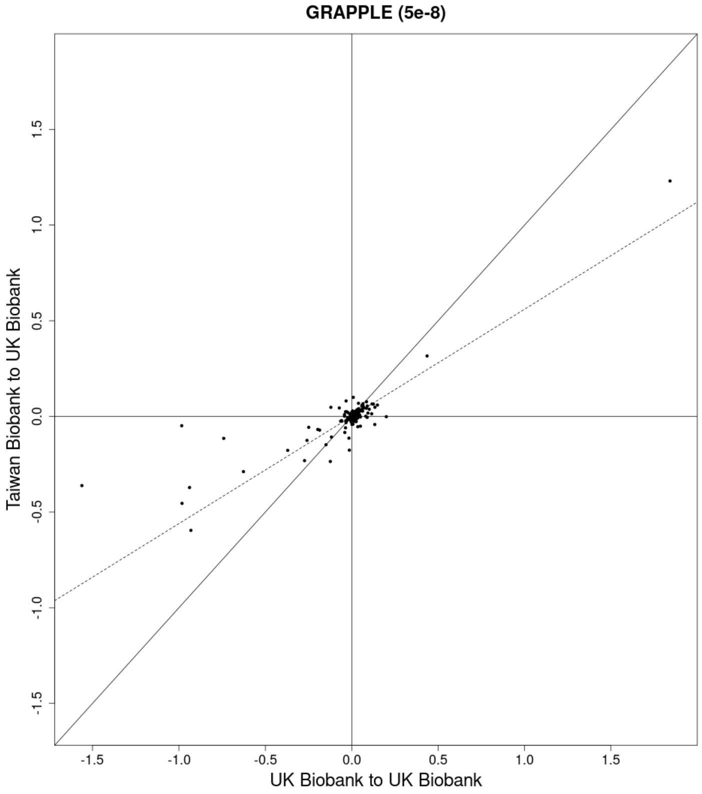
GRAPPLE (5e-8)
Taiwan Biobank to UK Biobank
UK Biobank to UK Biobank
-1.5
-1.0
-0.5
0.0
0.5
1.0
1.5

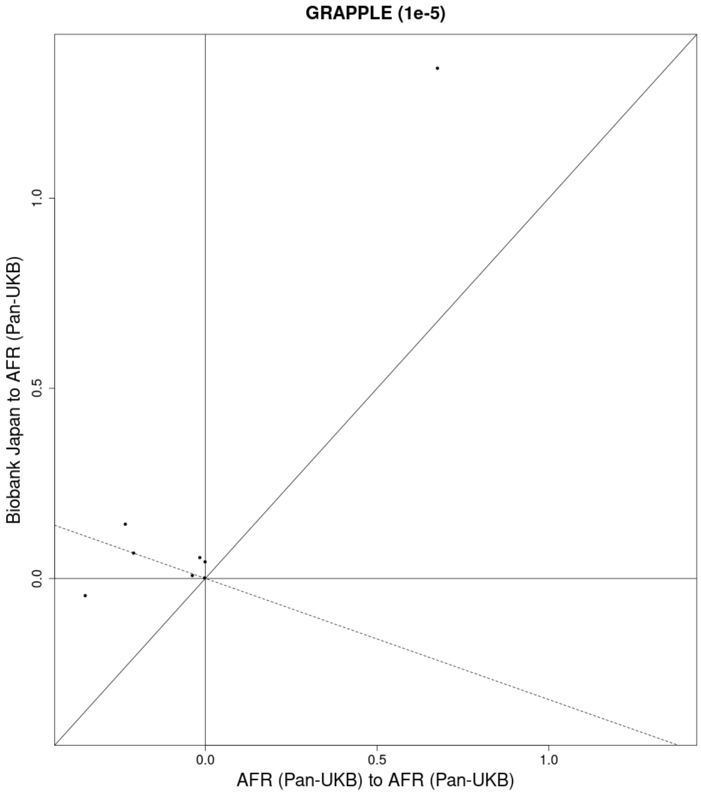
GRAPPLE (1e-5)
Biobank Japan to AFR (Pan-UKB)
AFR (Pan-UKB) to AFR (Pan-UKB)
0.0
0.5
1.0

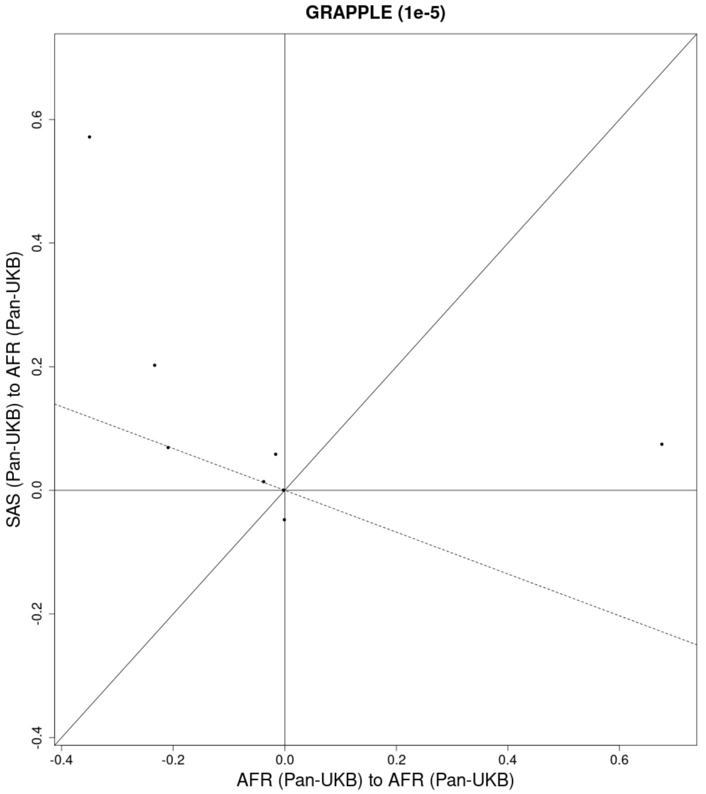


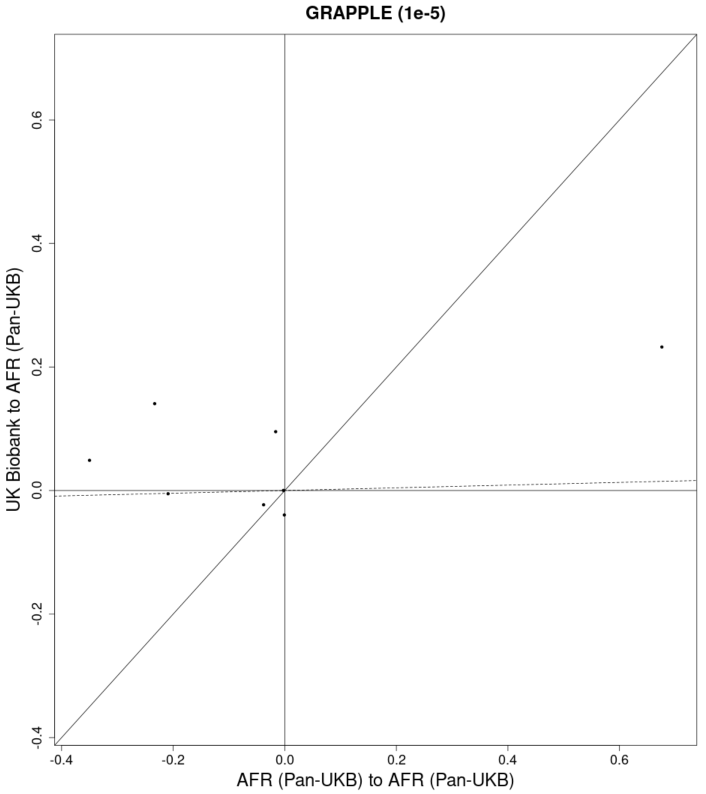


Figure S4: Cross-population vs. same-population MR estimates for 23 population pairs at both instrument selection thresholds and 3 population pairs - using BBJ, South Asian (Pan-UKB), and UKB studies as exposures for African (Pan-UKB) outcomes - at only an instrument selection threshold of $1 \times 10^{-5}$. All estimates were calculated using GRAPPLE. The dashed line represents the SIMEX-estimated shrinkage coefficient for the population pair and instrument selection threshold.

#### Supplemental References


[1] Joshua D. Angrist, Kathryn Graddy, and Guido W. Imbens. The Interpretation of Instrumental Variables Estimators in Simultaneous Equations Models with an Application to the Demand for Fish. *The Review of Economic Studies*, 67(3):499–527, July 2000. ISSN 0034-6527. doi: 10.1111/1467-937X.00141. URL `https://doi.org/10.1111/1467-937X.00141`.

[2] Stephen Burgess, Neil M. Davies, Simon G. Thompson, and EPIC-InterAct Consortium. Instrumental variable analysis with a nonlinear exposure-outcome relationship. *Epidemiology (Cambridge, Mass.)*, 25(6):877–885, November 2014. ISSN 1531-5487. doi: 10.1097/EDE.0000000000000161.

[3] Stephen Burgess and Jeremy A. Labrecque. Mendelian randomization with a binary exposure variable: interpretation and presentation of causal estimates. *European Journal of Epidemiology*, 33(10):947–952, 2018. ISSN 0393-2990. doi: 10.1007/s10654-018-0424-6. URL `https://pmc.ncbi.nlm.nih.gov/articles/PMC6153517/`.

[4] Eleanor Sanderson, M. Mario Glymour, Michael Holmes, Hyunseung Kang, Jean Morrison, Marcus Munafo, Tom Palmer, C. Mary Schooling, Chris Wallace, Qingyuan Zhao, and George Davey Smith. Mendelian randomization. *Nature Review Methods Primers*, 2(6), 2022.

[5] Fernando Pires Hartwig, Linbo Wang, George Davey Smith, and Neil Martin Davies. Average Causal Effect Estimation Via Instrumental Variables: the No Simultaneous Heterogeneity Assumption. *Epidemiology*, 34(3):325, May 2023. ISSN 1044-3983. doi: 10.1097/EDE.0000000000001596. URL `https://journals.lww.com/epidem/abstract/2023/05000/average_causal_effect_estimation_via_instrumental.4.aspx`.

[6] Armstrong. CHEBYSHEV INEQUALITIES AND COMONOTONICITY. *Real Analysis Exchange*, 19(1):266, 1993. ISSN 01471937. doi: 10.2307/44153837. URL `https://projecteuclid.org/journals/real-analysis-exchange/volume-19/issue-1/CHEBYSHEV-INEQUALITIES-AND-COMONOTONICITY/10.2307/44153837.pdf`.

[7] Gerhard Moser, Sang Hong Lee, Ben J. Hayes, Michael E. Goddard, Naomi R. Wray, and Peter M. Visscher. Simultaneous Discovery, Estimation and Prediction Analysis of Complex Traits Using a Bayesian Mixture Model. *PLoS Genetics*, 11 (4):e1004969, April 2015. ISSN 1553-7390. doi: 10.1371/journal.pgen.1004969. URL `https://pmc.ncbi.nlm.nih.gov/articles/PMC4388571/`.

[8] Luke J. O'Connor. The distribution of common-variant effect sizes. *Nature Genetics*, 53(8):1243–1249, August 2021. ISSN 1546-1718. doi: 10.1038/s41588-021-00901-3. URL `https://www.nature.com/articles/s41588-021-00901-3`. Publisher: Nature Publishing Group.

[9] Jian Zeng, Ronald de Vlaming, Yang Wu, Matthew R. Robinson, Luke R. Lloyd-Jones, Loic Yengo, Chloe X. Yap, Angli Xue, Julia Sidorenko, Allan F. McRae, Joseph E. Powell, Grant W. Montgomery, Andres Metspalu, Tonu Esko, Greg Gibson, Naomi R. Wray, Peter M. Visscher, and Jian Yang. Signatures of negative selection in the genetic architecture of human complex traits. *Nature Genetics*, 50(5):746–753, May 2018. ISSN 1546-1718. doi: 10.1038/s41588-018-0101-4. URL `https://www.nature.com/articles/s41588-018-0101-4`. Publisher: Nature Publishing Group.

[10] Noah Lorincz-Comi, Yihe Yang, Gen Li, and Xiaofeng Zhu. Mrbee: A bias-corrected multivariable mendelian randomization method. *HGG Advances*, 5(3), 2024.

[11] Jingshu Wang, Qingyuan Zhao, Jack Bowden, Gibran Hemani, George Davey Smith, Dylan S. Small, and Nancy R. Zhang. Causal inference for heritable phenotypic risk factors using heterogeneous genetic instruments. *PLoS Genetics*, 17 (6), 2021.

[12] Gibran Hemani, Jie Zheng, Benjamin Elsworth, Kaitlin H Wade, Valeriia Haberland, Denis Baird, Charles Laurin, Stephen Burgess, Jack Bowden, Ryan Langdon, Vanessa Y Tan, James Yarmolinsky, Hashem A Shihab, Nicholas J Timpson, David M Evans, Caroline Relton, Richard M Martin, George Davey Smith, Tom R Gaunt, and Philip C Haycock. The mr-base platform supports systematic causal inference across the human phenome. *eLife*, 7, 2018.

[13] Gibran Hemani, Kate Tilling, and George Davey Smith. Orienting the causal relationship between imprecisely measured traits using gwas summary data. *PLoS Genetics*, 13(12), 2017.

[14] Sang Hong Lee, Michael E Goddard, Naomi R Wray, and Peter M Visscher. A better coefficient of determination for genetic profile analysis. *Genetic Epidemiology*, 36(3), 2012.

[15] J.R. Cook and L.A. Stefanski. Simulation-extrapolation estimation in parametric measurement error models. *Journal of the American Statistical Association*, 89(428):1314–1328, 1994.

[16] Yi Liu, Benjamin Elsworth, Pau Erola, Valeriia Haberland, Gibran Hemani, Matt Lyon, Jie Zheng, Oliver Lloyd, Marina Vabistsevits, and Tom R Gaunt. Epigraphdb: a database and data mining platform for health data science. *Bioinformatics*, 37(9):1304–1311, 2021.
